# Personalized HLA-aware TCR measure predicts the risk of symptomatic SARS-CoV-2 infection following booster vaccination

**DOI:** 10.64898/2026.11.10.26367219

**Authors:** Koshlan Mayer-Blackwell, Khiyam Hussain, Narges Ebrahimi, Emily S. Ford, Adam P. Dale, Jennifer Alderson, Sagida Bibi, Alasdair P.S. Munro, Saul N. Faust, Xinxue Liu, Teresa Lambe, Merryn Voysey, Andrew Fiore-Gartland

**Affiliations:** Vaccine and Infectious Disease Division, Fred Hutchinson Cancer Center, Seattle, USA; Vaccine Immunology Statistical Center, Fred Hutchinson Cancer Center, Seattle, USA; Oxford Vaccine Group, Department of Paediatrics, University of Oxford, Oxford, UK; Department of Medicine, University of Washington, Seattle, USA; NIHR Southampton Clinical Research Facility and Biomedical Research Centre, University Hospital Southampton NHS Foundation Trust; and Faculty of Medicine and Institute for Life Sciences, University of Southampton, Southampton, UK; Pandemic Sciences Institute (PSI), University of Oxford, Oxford, UK; Chinese Academy of Medical Sciences Oxford Institute, Oxford, UK

## Abstract

HLA-driven heterogeneity and limitations of live T cell functional assays routinely thwart assessment of cell-mediated immunity in vaccination trials. To address this, we developed a T-cell receptor (TCR)-based measure of immunogen recognition and evaluated it in COV-BOOST, a third-dose COVID-19 vaccine trial, during the Delta and Omicron waves (2021-2022, United Kingdom). We estimated HLA-aware Spike-reactive T cell (HA-T_S_) breadth from participant HLA genotypes, TCRβ repertoires, and a novel SARS-CoV-2 Spike HLA-linked reference catalog. Higher post-boost HA-T_S_ breadth was associated with lower symptomatic infection risk (HR 0.58, 95% CI 0.39–0.86). This association persisted after adjustment for neutralizing antibody titers and IFN-γ responses. Examination of HA-T_S_ across prior studies showed that it reflects participant genotype, exposure intensity, and time from exposure to sampling. These findings highlight the possibility that estimates of antigen-specific T cell repertoire breadth could serve as a correlate of protection to accelerate vaccine development.

## Introduction

Immunological memory from infection and vaccination has reduced the population-level burden of severe COVID-19, even as reinfection waves drive viral escape from common antibody neutralization sites ^1–3^. Meta-analyses of vaccine efficacy show neutralizing antibodies (nAbs) correlate strongly with protection against early post-vaccination infections ^4–7^. However, this humoral protection wanes over time, is strain-dependent, and may not fully explain observed levels of vaccine efficacy ^6,8,9^. Thus, T cell mediated immunity has been proposed as a complementary mechanism underlying lasting protection against disease progression and severe clinical outcomes ^10–12^.

Considerable evidence supports a role for cellular immunity in the control of SARS-CoV-2 infection in animal models and humans. In B cell-deficient mice, T cells were sufficient to provide antibody-independent protection against severe SARS-CoV-2 disease ^13,14^, and robust anti-SARS-CoV-2 T cell responses have also been observed in humans with B cell immunodeficiencies ^15^. In pediatric cohorts, the magnitude of spike-reactive IFN-γ- and IL-2-producing T cells following vaccination has been inversely associated with the risk of symptomatic infection over subsequent months ^16^, and a household exposure study reported that early induction of SARS-CoV-2-specific IFN-γ-secreting T cells was associated with reduced viral load and milder disease ^17^. Although T cell responses were not directly analyzed, an analysis of humoral correlates of protection in a phase 3 trial of an adenovirus vectored vaccine (AZD1222), observed substantial vaccine efficacy even among participants with low levels of measured neutralizing antibodies ^6,7^. Recently, an examination of SARS-CoV-2-naïve individuals receiving a single dose of Ad26.COV2.S vaccine early in the pandemic showed that a higher fraction of TCRs associated with SARS-CoV-2 Spike antigen, measured 29 days post-vaccination, was associated with a reduced risk of severe, but not mild, COVID-19 during a 200 day follow-up ^18^.

Despite these lines of evidence for the protective role of T cells against SARS-CoV-2, few human studies have directly linked features of vaccine-elicited T cell repertoires to subsequent risk of SARS-CoV-2 infection using longitudinal designs. Assessment of vaccine-induced cellular immunity in large trials is complicated by HLA-driven heterogeneity in T cell responses, the cost of cryopreservation, and lab-to-lab variability in live-cell assays. To address this, we developed an HLA-aware (HA) analysis of T cell receptor (TCR) sequencing to estimate the clonotypic breadth of immunogen recognition. The approach utilizes a newly constructed catalog of spike-specific TCRs developed from public data sources, to tally matching TCRs from each participant’s repertoire based on their HLA genotype. Applying this approach to a subcohort of the COV-BOOST trial ^19,20^, we investigated the association between TCRβ repertoire diversity and the risk of symptomatic SARS-CoV-2 breakthrough infection. The subcohort comprised SARS-CoV-2-naive participants who had previously received two doses of either an mRNA or ChAdOx COVID-19 vaccines. Participants subsequently received one of several booster vaccine regimens. Here we report that greater diversity of spike-reactive TCRs in peripheral blood TCRβ repertoires was associated with a lower risk of symptomatic SARS-CoV-2 infection over the nine-month follow-up period.

## Results

### Booster vaccination increases breadth of antigen-specific TCRs

In light of the challenges in storing PBMCs and standardizing live-cell functional assays for assessing vaccine specific T cell responses in large clinical trials, we developed an HLA-aware TCRβ sequencing-based approach to estimate the breadth of immunogen recognition by SARS-CoV-2 spike-reactive T cells (T_S_). This metric, which we term HA-T_S_ breadth, assesses the proportion of spike-specific T cells in a repertoire by tabulating weighted matches to an HLA-defined SARS-CoV-2 spike TCRβ catalog (see Methods) and dividing the number of matches by total number of unique clonotypes. High resolution HLA genotypes from each participant were incorporated to compute participant-level, HLA-matched T_S_ breadth scores by querying a catalog of thousands of TCR sequences with established experimental or statistical restriction to specific HLA alleles (i.e., HLA-A, -B, -C, or -DRB1) or functional heterodimers (i.e., DQA1-DQB1 / DPA1-DPB1) (see Methods, Extended Data **Fig. 1**). Briefly, each HLA-compatible TCRβ match is assigned a weight between 0 and 1 to reflect the uncertainty in sequence-based matching relative to the TCRβ’s baseline prevalence in HLA-matched SARS-CoV-2 naive repertoires; a weight close to 1 indicates high certainty that the detected TCRβ is spike-specific in that HLA-context, evidenced by its low prevalence in HLA-matched SARS-CoV-2 naive repertoires (see Methods). We computed HA-T_S_ breadth in paired pre-(Day 0) and post-boost (Day 14) repertoire samples for a subset of participants within the COV-BOOST trial (n = 120). As expected, booster administration induced a significant increase in the median HA-T_S_ breadth score, increasing from 1.9 ✕ 10^-4^ (IQR: 1.1 ✕ 10^-4^ – 2.4 ✕ 10^-4^) pre-boost to 5.0 ✕ 10^-4^ (IQR: 3.6 ✕ 10^-4^ – 7.5 ✕ 10^-4^) post-boost (p < 0.0001, Wilcoxon signed-rank test, Extended Data **Fig. 2A**). Components of HA-T_S_ breadth attributable to CD4^+^ and CD8^+^ receptors both significantly increased between pre-and post-boost repertoires, with a moderate rank correlation (ρ = 0.55, p < 0.0001) between the absolute participant-level changes in CD4^+^ and CD8^+^ T_S_ breadth from Day 0 to Day 14 (Extended Data **Fig. 2B-D**).

### Spike-reactive T cell breadth is associated with reduced risk of symptomatic SARS-CoV-2 infection

To examine whether HA-T_S_ breadth was associated with subsequent risk of symptomatic SARS-CoV-2 infection within the COV-BOOST trial, we fit covariate-adjusted inverse probability–weighted Cox proportional hazard models, stratifying by vaccination start month to account for calendar effects (June 2021 vs. November 2021; **Fig. 1A-B**). In these models, participants with high HA-T_S_ breadth (defined by a median split of Day 14 values) had an estimated hazard of symptomatic infection that was approximately one-third that of participants with low HA-T_S_ breadth (HR = 0.32; 95% CI 0.16–0.67; p = 0.002; **Fig. 1E**, model 1). When modeled as a continuous variable, a doubling of HA-T_S_ breadth was associated with a 42% reduction in the risk of symptomatic infection (HR = 0.58; 95% CI 0.39–0.86; p = 0.007; **Fig. 1E**, model 2).

**Fig. 1:**
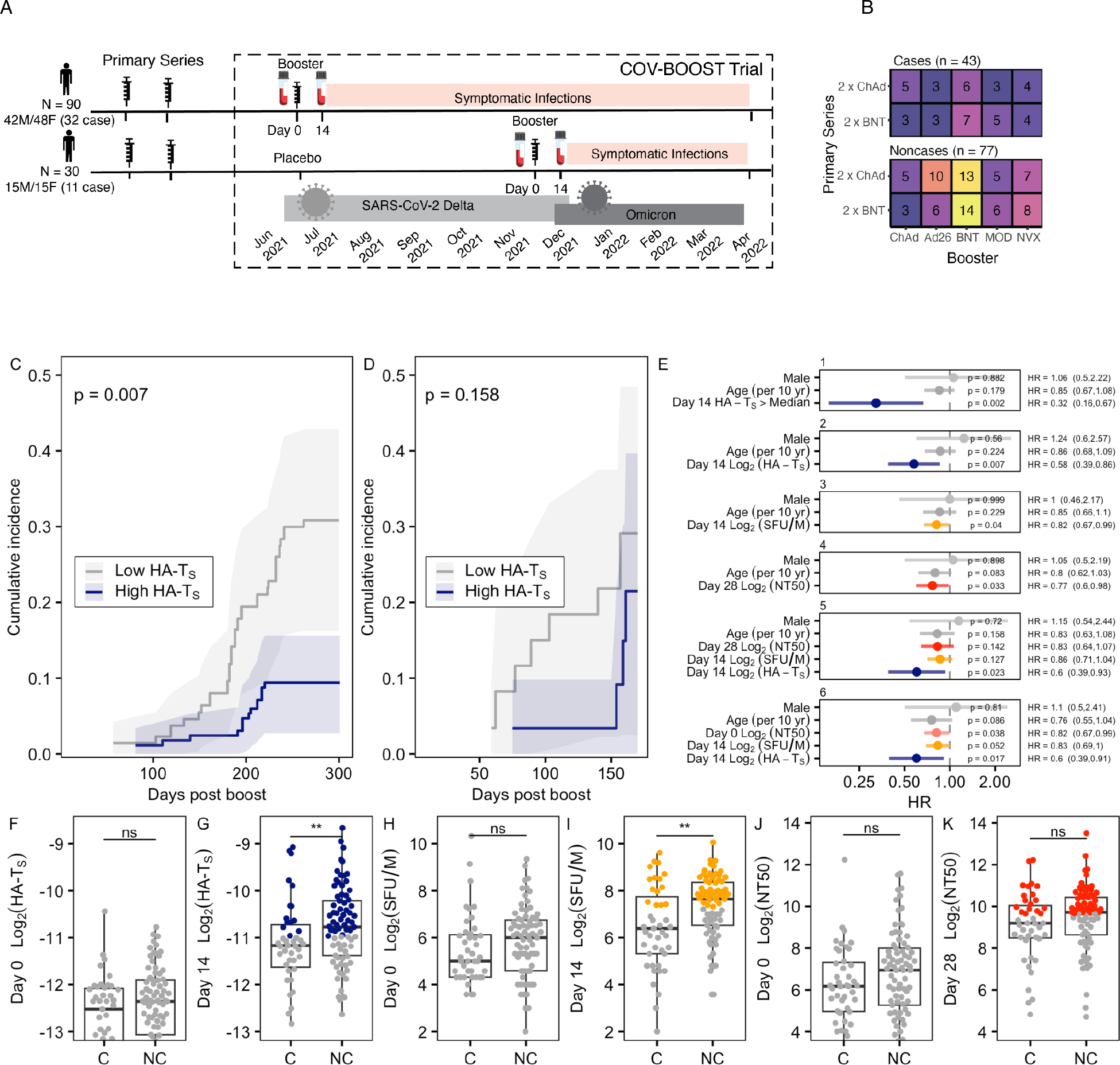
Spike-reactive T-cell breadth is negatively associated with symptomatic SARS-CoV-2 infection. (**A**) Timeline of follow-up after vaccination and blood collection for TCRβ sequencing, antibody neutralization, ELISpot assays. Participants were vaccinated in June 2021 or November 2021 as part of COV-BOOST clinical trial.(**B**) Number of analyzed participants by case status and vaccine series. Booster vaccines included: adenoviral (ChAdOx1, Ad26), mRNA (BNT162b2, mRNA-1273), or protein subunit (NVX-CoV2373).(**C–D**) Kaplan–Meier estimates of cumulative incidence of symptomatic SARS-CoV-2 infection during follow-up split by study-wide median split of Day 14 HA-T_S_ breadth, shown for groups receiving booster vaccination in June 2021 (n = 90) or November 2021 (n = 30) booster vaccination. P-values are from a log-rank test. (**E**) Hazard ratios from covariate-adjusted inverse probability–weighted proportional hazards models including HA-T_S_ breadth, neutralizing antibody titers (NT50), and/or spike-specific IFNγ ELISpot responses. (**F-K**) T_S_ breadth, ELISpot responses (SFU/M), and antibody potency (NT50) at baseline (Day 0) and post-boost (Day 14 or Day 28) in symptomatic cases (C) and noncases (NC), with p-values based on Wilcoxon rank-sum test. Colored points represent values above Day 14 median value corresponding with Kaplan-Meier plots in Extended Data Fig. 3. In panel F, only values greater than Log_2_(HA-T_S_) > −13 and Log_2_(NT50) > 4 are shown so that the axes are comparable between panels.

Supporting these models, cumulative incidence curves for participants with high vs. low HA-T_S_ breadth showed differences in incidence during both the June 2021 and November 2021 vaccination periods (**Fig. 1C-D**). There was evidence for a reduced hazard of symptomatic infection among participants with high HA-T_S_ breadth in the June cohort (log-rank test, p = 0.007 [n = 90]), with a similar, but non-significant trend present in the smaller November cohort (p = 0.158 [n = 30]). We also observed that pre-boost (Day 0) HA-T_S_ breadth tended to be higher among participants in the sample who did not go on to experience a symptomatic infection compared with those who did, although the difference was not statistically significant (Wilcoxon rank-sum test, p = 0.12; **Fig. 1F**). By Day 14 post-boost, this separation was more pronounced, with HA-T_S_ breadth significantly higher among non-cases (p = 0.0096; **Fig. 1G**), suggesting that booster dose may directly diversify the spike reactive T cell repertoire or at least expand pre-existing memory T cells to a frequency robustly detected by the TCRseq assay.

Next, we examined how Day 14 HA-T_S_ breadth related to other measures of post-vaccine immunity. Compared with cases, non-cases tended to have higher spike-specific IFN-γ responses measured by ELISpot (SFU/M **Fig. 1H–I)**, but the two groups had similar neutralizing antibody titers (NT50; **Fig. 1J–K**). HA-T_S_ breadth showed a modest positive rank correlation with ELISpot responses (ρ = 0.22, p = 0.01; Extended Data **Fig. 3**), but little correlation with NT_50_ values (ρ = 0.15, p = 0.10; Extended Data **Fig. 3**). To better understand the correlation between HA-T_S_ and IFN-γ responses, we examined correlations between TCRseq metrics – broken down by CD4^+^ and CD8^+^ features – and ELISpot levels (Extended Data **Fig. 4**). We observed that receptors linked to class I HLA alleles (HA-T_S,CD8+_) were more strongly correlated with ELISpot than receptors linked to class II HLA alleles (HA-T_S,CD4+_). We further considered the clonal abundance of spike reactive T cells, termed here as HA-T_S_ depth, computed as the weighted sum of clonal frequencies receptors matching the HLA-defined SARS-CoV-2 spike TCRβ catalog (see methods). The diversity (HA-T_S,CD8+_ breadth), abundance (HA-T_S,CD8+_ depth), and change in both metrics between Day 0 and Day 14, were all significantly correlated with IFN-γ functional responses (Extended Data **Fig. 4A-D**). Neither HA-T_S,CD4+_ breadth or depth showed a significant correlation with functional IFN-γ responses (Extended Data **Fig. 4E-G**).

When overall HA-T_S_ breadth was included alongside NT_50_, ELISpot, or both, in covariate-adjusted proportional hazards models, the estimated association between HA-T_S_ breadth and symptomatic infection risk remained consistent across models (**Fig. 1E** models 2, 5, and 6). In models including only log2-transformed ELISpot (model 3) or NT50 (model 4), controlling for age and sex, higher values of each measure were independently associated with lower hazard of symptomatic infection (ELISpot: HR = 0.82, 95% CI 0.67–0.99, p = 0.04; NT50: HR = 0.77, 95% CI 0.60–0.98, p = 0.03). Notably, although antibody neutralization titres increased following vaccination in most participants, the pre-boost NT_50_ values were more strongly associated with reduced infection risk compared with the Day 28 NT_50_ values (**Fig. 1E**, models 5 and 6, Extended Data **Fig. 3**). Adjustment for pre-boost (Day 0) NT_50_ did not meaningfully alter the estimated association between HA-T_S_ breadth and symptomatic infection risk (**Fig. 1E**, model 6). Multiple sensitivity analyses confirmed that the association between HA-T_S_ and symptomatic breakthrough risk was robust to the non-random processing of case and noncase samples across DNA extraction batches, including analyses restricted to deeply-sequenced samples and analyses adjusting for total recovered repertoire diversity after resampling (Methods, Extended Data **Fig. 5**). Taken together, these primary and sensitivity analyses indicate that estimates of spike-reactive clonotypic breadth (i.e., the diversity of antigen-specific T cells) captures a dimension of vaccine-enhanced immunity in this cohort that is distinct from, and potentially complementary to, peak neutralizing antibody potency and the magnitude of IFN-γ T-cell responses.

### HLA-aware T_S_ breadth outperforms HLA-agnostic measures in resolving breakthrough cases

To compare our newly developed HLA-aware T cell breadth measure to previously published HLA-agnostic TCRseq-based metrics of Spike immunogen recognition ^21–23^, we evaluated both types of metric at baseline (Day 0) and post-vaccination (Day 14) in the same participants, stratified by breakthrough infection status (case vs. non-case). The HA-T_S_ breadth measure did not significantly differ between cases and non-cases at Day 0 in aggregate or by CD4+ versus CD8+ compartment (Extended Data **Fig. 6A,C,E**; overall p = 0.12; CD4+ p = 0.11; CD8+ p = 0.13), but by Day 14 non-cases showed significantly greater breadth than cases in aggregate and separately by CD4+ versus CD8+ compartments (Extended Data **Fig. 6B, D, F**; overall p = 0.0096; CD4+ p = 0.022; CD8+ p = 0.021).

In contrast, the analogous HLA-agnostic Adaptive Biotechnologies Spike breadth metric did not as well separate cases from non-cases at either timepoint in aggregate or by T cell subset (all p ≥ 0.10), with the strongest but still non-significant signal in Day 14 CD4+ breadth (p = 0.096). This absence of discrimination extended to the other Adaptive HLA-agnostic metrics – including MIRA-derived breadth, the Spike version 3 (V3) classifier score, and Spike score. None of the HLA-agnostic TCR metrics robustly separated cases from non-cases at either timepoint (Extended Data **Fig. 6G-L,S-X**, all p ≥ 0.059).

At the per-participant level, HLA-aware and HLA-agnostic breadth showed a modest positive rank correlation. Concordance between metrics was strongest for the overall (combined CD4^+^ and CD8^+^) and CD4^+^ compartments (ρ = 0.34 and 0.43 at Day 0 and Day 14, respectively; CD4+ ρ = 0.25 and 0.33) and weakest for CD8^+^ (ρ = 0.15 and 0.19 at Day 0 and Day 14, respectively). Across all metric pairs, per-sample comparisons fell predominantly above the 1:1 line (Extended Data **Fig. 6M–R**), indicating that the HLA-aware assay captured a greater total number of sequences than the previous HLA-agnostic measures. Many samples had non-detectable CD8^+^ S breadth by the prior HLA-agnostic Adaptive method (Extended Data **Fig. 6 K,L**).

These results indicate that the newly developed HA-T_S_ metrics capture a vaccine-responsive signal associated with reduced risk of breakthrough infection that was not as well resolved by previously published metrics. Although the strong positive correlation seen in Day 14 between overall HA-T_S_ and Adaptive Biotechnologies’ Spike breadth metric (Extended Data **Fig. 6N**) suggests a shared signal captured by both methods. The prior HLA-agnostic metric were constructed for classification of exposure states and likely selected a smaller set of diagnostic “enhanced sequences” that were highly enriched in post COVID-19 repertoires ^21^. The greater sensitivity to detect more clonotypes via HA-T_S_ likely reflects two aspects of our approach. First, the new HLA-aware SARS-CoV-2 Spike catalog draws on experimental data from post-vaccine repertoires, which may be favorable for identifying Spike-specific responses given lack of competition with T cell responses to other viral antigens (e.g., epitopes in nucleocapsid protein or ORF1) seen in natural infection; second, our approach accommodates more ambiguous, higher probability of V(D)J-recombination sequences via confidence weighting and tabulates catalog hits only in participants carrying the corresponding HLA, potentially allowing more Spike-specific sequences to be included in the reference set.

### HLA-aware T_S_ breadth reflects varied SARS-CoV-2 exposures, time since infection, and disease severities

To contextualize booster vaccination with other SARS-CoV-2 exposures, we imputed participants HLA genotypes directly from their TCRβ repertoires using the TCR2HLA software ^24^ and estimated HA-T_S_ breadth in TCR repertoires collected in prior studies of acute COVID-19 ^25^, convalescence ^22^, and vaccinated individuals ^26,27^, as well as pre-pandemic SARS-CoV-2 naive repertoires ^28^. The breadth of spike-associated receptors in SARS-CoV-2 naive repertoires collected before 2019 was heterogeneous, but the distribution of HA-T_S_ breadth values in the naive repertoires was lower than in repertoires following any mode of SARS-CoV-2 S exposure (**Fig. 2A**). In SARS-CoV-2 naive repertoires, we detected no substantial difference in T_S_ breadth in repertoires from CMV seropositive versus seronegative individuals (**Fig. 2A).** Prior to booster vaccination (Day 0, COV-BOOST), HA-T_S_ breadth levels from previously vaccinated participants were only slightly elevated (p < 0.0001) compared to naive repertoires, consistent with the contraction of T cell memory from the earlier vaccination. Post-boost levels were similar to those observed in another study where healthy participants received two doses of mRNA vaccine (vs. COV3006; p = 0.54) but higher than in repertoires from prior studies where participants received 1-2 doses of ChAdOx (vs. COV002; p= 0.026) or 1 dose of Ad26 (vs. ENSEMBLE; p < 0.0001) (**Fig. 2A**).

**Fig. 2:**
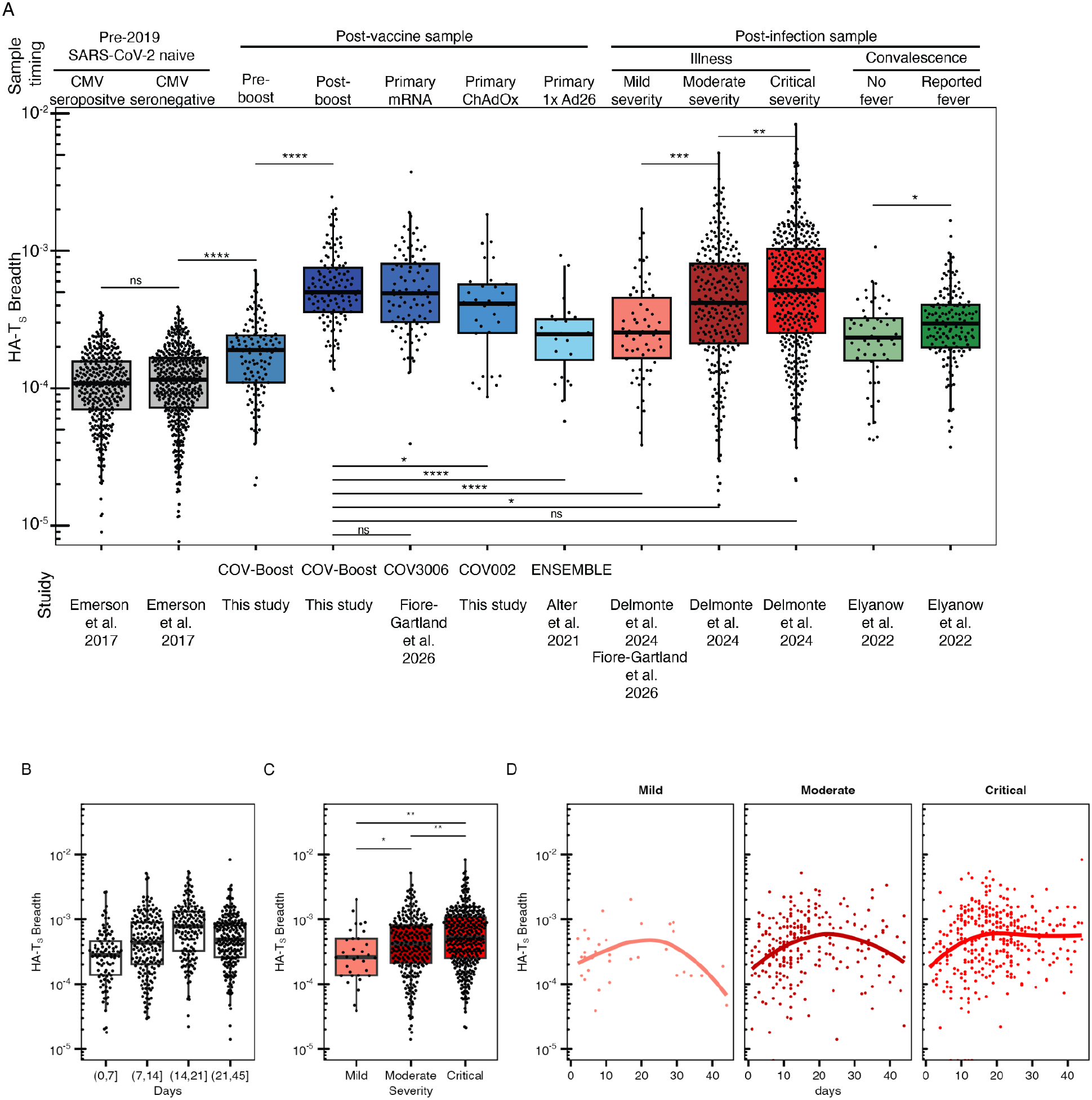
Spike-reactive T-cell breadth reflects exposure severity and time from last exposure. (**A**) Distribution of HLA-aware spike-reactive clonotypic breadth (HA-T_S_) across distinct exposure cohorts, including naive repertoires (CMV seropositive/negative, gray), vaccination groups (blue), acute infections across varying levels of severity (red), and convalescence (green). Horizontal brackets indicate pairwise statistical comparisons with p-values (Wilcoxon rank-sum test). Mild infection group combines persons with 45 days of symptoms (Delmonte et al. study) or 45 days of PCR confirmed infection (Fiore Gartand et al. study). (**B**) Analysis of a post-infection cohort (Delmonte et al.) showing T_S_ binned by days from symptom onset. Brackets denote statistical significance between defined time intervals. (Wilcoxon rank-sum test). (**C**) Comparison of HA-T_S_ within the acute infection cohort, stratified by overall disease severity (Mild, Moderate, and Critical) in the Delmonte cohort. (**D**) Longitudinal tracking of HA-T_S_ versus time (days post-symptom onset), faceted by disease severity. Each person was sampled once, with individual data points representing sample timing. Solid lines indicate LOESS smoothed trend lines. Statistical significance levels are indicated by asterisks: ****, p < 0.0001; ***, p <0.001; **, p <0.01; *, p < 0.05; ns, not significant.

Across two prior studies of natural infection, with repertoires sampled during or shortly after acute infection, HA-T_S_ breadth was associated with reported disease severity, with the highest HA-T_S_ values observed in critical illness. (**Fig. 2A**). HA-T_S_ breadth also appeared sensitive to post-infection sample timing with higher HA-T_S_ breadth observed in the third week after symptom onset (days 14-21) compared with earlier and later timepoints (**Fig. 2B**). Similar kinetics trends were observed across severity groups (**Fig. 2C,D**) with splines suggesting critical cases resulted in greater or longer lasting HA-T_S_ breadth **(Fig. 2D**). Consistent with this, in persons recovered from COVID-19, HA-T_S_ breadth in convalescent repertoires collected within 90 days of initial symptoms (Elyanow et al. 2022) were considerably lower than those in acute phase patients (Delmonte et al. 2024), but levels were elevated among those who had experienced a fever compared with those who had not (**Fig. 2A**). The capacity to use HA-T_S_ breadth to discriminate naive individuals from those with disease varied considerably by disease severity, with the highest separation observed for critical (AUC = 0.917) and moderate cases (AUC = 0.880), and lower performance for convalescent repertoires with fever (AUC = 0.866) or mild (AUC = 0.8) infection. For a practical comparison, Day 14 T_S_ levels measured post-boost in the COV-Boost cohort were similar to the diversity of spike-specific memory observed in persons who had experienced a severe or critical COVID-19 infection (**Fig. 2A**).

Within the COV-Boost cohort, we examined whether HA-T_S_ breadth varied by either the primary or boost vaccine modality. Day 0 HA-T_S_ did not differ significantly between participants who had received the mRNA or ChAdOx primary vaccine series (Extended Data **Fig. 7A**). Median Day 14 HA-T_S_ and fold change in HA-T_S_ was lowest among participants who only received ChAdOx (ChAdOx(3)), but difference in HA-T_S_ did not reach statistical significance between ChAdOx(3) and the other booster modalities (Extended Data **Fig. 7A**). Participants who received ChAdOx, as primary and booster (ChAdOx(3)), did have lower fold change in NT50 (Extended Data **Fig. 7B**) and lower fold change ELISpot (Extended Data **Fig. 7C**) responses compared with the group that received either a boost of Ad26 or ChAdOx after previously receiving an mRNA primary vaccination. An attenuated immune response is consistent with prior studies that show mechanistic inhibition of vaccine response to homologous ChAdOx vectored re-vaccination by pre-existing anti-ChAdOx serum antibodies ^29–31^.

### Expected HLA-intrinsic breadth, based on HLA genotype alone, partially predicts clinical risk

We defined HA-T_S_ breadth as the proportion of unique TCRs in a repertoire expected to recognize the spike antigen. Because this metric relies on matching TCRs to a subset of the reference catalog defined by an individual’s HLA genotype, the maximum measurable breadth is inherently linked to each person’s specific HLA alleles and their catalog representation. While the HLA-defined catalog may reflect biases in its curation, we hypothesized that the differences in the potential breadth scores associated with combinations of HLA alleles may reflect genuine biological differences in the presentation efficiency of distinct HLA proteins. That is, if specific HLA variants poorly present vaccine or viral peptides, individuals carrying more weak-presenting alleles may mount a weaker T cell response overall and potentially face higher clinical risk. It is also possible that the composition of HLA-defined catalogs could be influenced by observational bias. For instance, highly prevalent HLA alleles afford greater statistical power for discovering public TCRs ^32^, and experimental techniques like tetramer staining inherently bias libraries toward known class I presented epitopes.

To address participant-level differences in HLA-SARS-CoV-2 S catalog overlap, we attempted to capture the potential influence of HLA genotype on HA-Ts breadth, without utilizing an individual’s TCR repertoire (Extended Data **Fig. 8**). First we consider an individual’s expected HLA-intrinsic Breadth (E[HA-T_S,HLA_]), which we simulated based on a participant’s complement of class I (HLA-A, B, C) and class II (DRB1, DQA/B, DPA/B) alleles and the analogous allele-specific T_S_ breadth contributions observed in independent cohort of individuals with a defined SARS-CoV-2 S vaccine exposure. For each individual and their HLA genotype, we used Monte Carlo sampling across the relevant allele-specific components to simulate a potential distribution of expected breadth scores; we used the median score to represent their expected HLA-intrinsic spike reactive breadth E[HA-T_S,HLA_]. We can then compute a residual breadth ratio (HA-T_S_/ E[HA-T_S,HLA_]), which is the ratio of observed to expected breadth; conceptually this captures whether an individual over- or under-performed the genetic baseline (Extended Data **Fig. 8**). Similarly, we can compute a breadth quantile score, defined as the percentile rank of each individual’s observed breadth relative to the full distribution of expected breadth scores from the Monte Carlo sampling.

### Expected HLA-intrinsic T_S_ breadth is associated with lower risk of symptomatic COVID-19

We used TCR repertoires, with TCR2HLA imputed HLA genotypes, from a previously published study (COVPN3006; repertoires sampled 15 to 45 days post second dose mRNA vaccination), to sample per-allele breadth contributions and compute the expected HLA-intrinsic breadth E[T_S,HLA_] for each participant in the primary COV-BOOST cohort (**Fig. 3A**). We found that the expected HLA-intrinsic breadth was significantly lower among individuals who developed a symptomatic breakthrough infection compared to non-cases (**Fig. 3B**, p = 0.02), mirroring the differences seen in the directly observed HA-T_S_ breadth (**Fig. 3C**, p = 0.009). The residual ratio HA-T_S_/ E[HA-T_S,HLA_] trended lower in cases, though this difference was not statistically significant (**Fig. 3D**), and a similar pattern emerged for the breadth quantile scores (**Fig. 3E**). Together, these findings strongly suggest that a participant’s underlying HLA genotype and the SARS-CoV-2 S catalog composition were dominant drivers of the large differences observed in post-boost T cell breadth, in addition to factors related to vaccine regimen or individual immunity playing a minor role. Interestingly, despite a moderately strong rank correlation between E[HA-T_S,HLA_] and observed HA-T_S_ (**Fig. 3**F, rho = 0.57, p < 0.0001), the expected breadth was not significantly correlated with Day 14 ELISpot SFU/M (**Fig. 3G**, rho = 0.047, p = 0.61) versus the residual breadth ratio HA-T_S_/ E[HA-T_S,HLA_] (**Fig. 3**H, rho = 0.24, p = 0.0087) which was. Thus, while HLA genetics may strongly predict the general capacity for a diverse pool of antigen-specific cells, other immunological variation that promote the expansion of these cells may also enhance effector capacity measured at Day 14.

**Fig 3:**
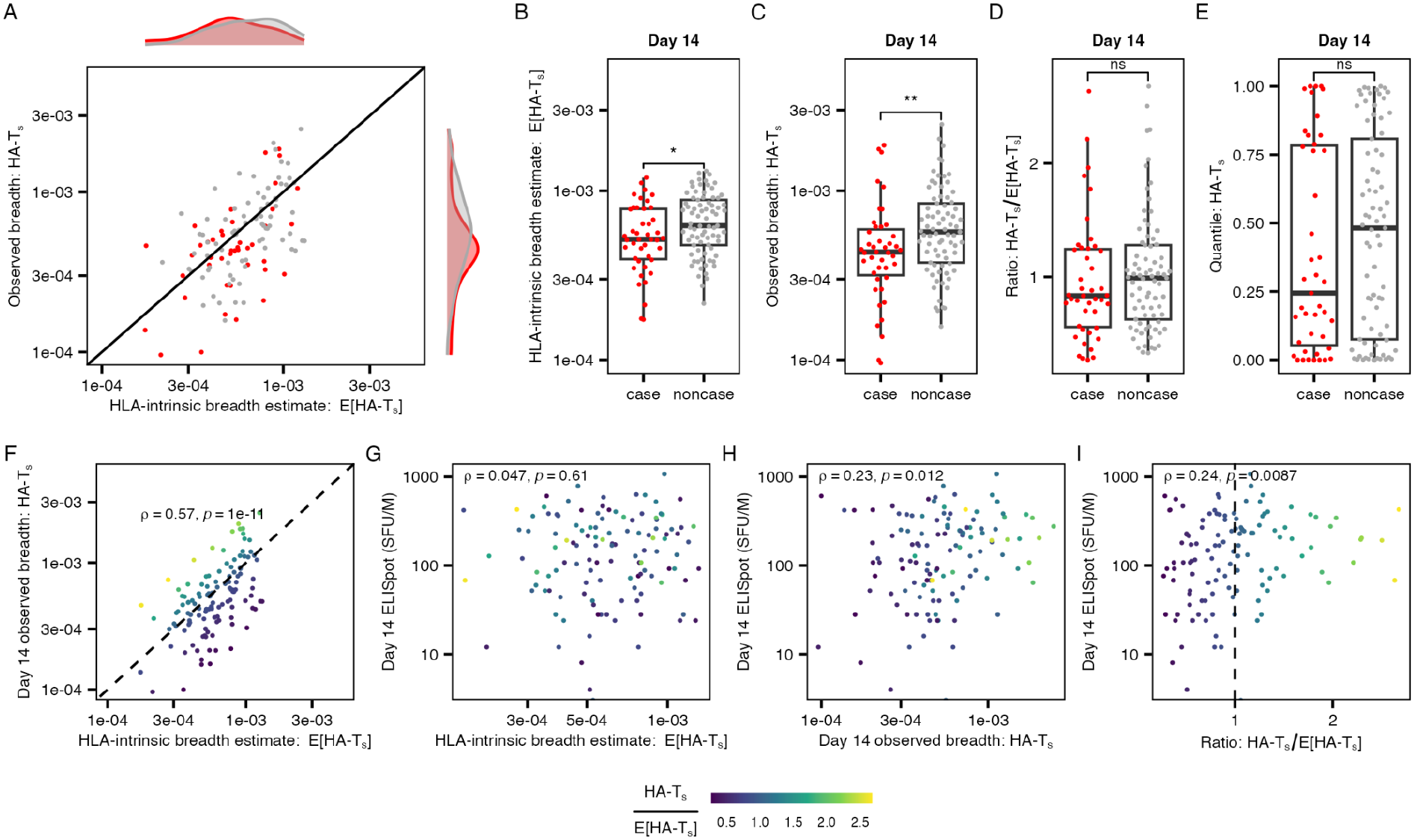
Expected HLA-intrinsic breadth predicts risk of symptomatic infection. **(A)** Expected HLA-intrinsic breadth (E[HA-T_S_]) versus observed (HA-T_S_) breadth for cases (red) and noncases (gray). **(B–E)** Distribution across cases versus noncases for (B) E[HA-T-_S_], (C) HA-T_S_, (D) HA-T_S_/E[HA-T_S_] ratio, and (E) HA-T_S_ quantile. (**F**) Correlation between E[HA-T_S_] and HA-T_S_ with point color showing the HA-T_S_/E/[HA-T_S_] ratio (**G–I)** Correlation of Day 14 ELISpot SFU/M with (**G**) E[HA-T_S_], (**H**) HA-T_S_, and (I) HA-T_S_/E/[HA-T_S_] ratio. Colors are consistent across panels F-I. **, p <0.01; *, p < 0.05, ns, not significant.

### Expected HLA-intrinsic T_S_ Breadth and Severe Disease

Next, we investigated whether expected HLA-intrinsic breadth could identify individuals at high risk for severe clinical outcomes following primary infection. Because the breakthrough cases in the COV-BOOST subcohort were mild, we examined participants in the Delmonte et al. study ^25^, which collected repertoires from unvaccinated individuals after infection, including many with severe illness. We expected that severe disease would drive the clonal expansion of a highly diverse spike-reactive TCR repertoire during the acute and convalescent phases due to prolonged antigenic load. Indeed, following infection, we observed maximal HA-T_S_ breadth in patients with critical COVID-19 compared to those with moderate or mild symptoms (**Fig. 2**C). To assess if this increase was driven by exposure intensity rather than underlying antigen presentation potential, we calculated each participant’s expected HLA-intrinsic breadth (**Fig. 4**). Notably, we found higher E[HA-T_S,HLA_] in people with mild disease (p = 0.033), but no systematic difference in E[HA-T_S,HLA_] between patients with moderate versus critically severe disease (**Fig. 4A**). Accounting for likely antigen exposure strength, the HA-T_S_/ E[HA-T_S,HLA_] ratio was elevated in critical vs. moderate cases (**Fig. 4**B, p =0.0006).

**Fig 4:**
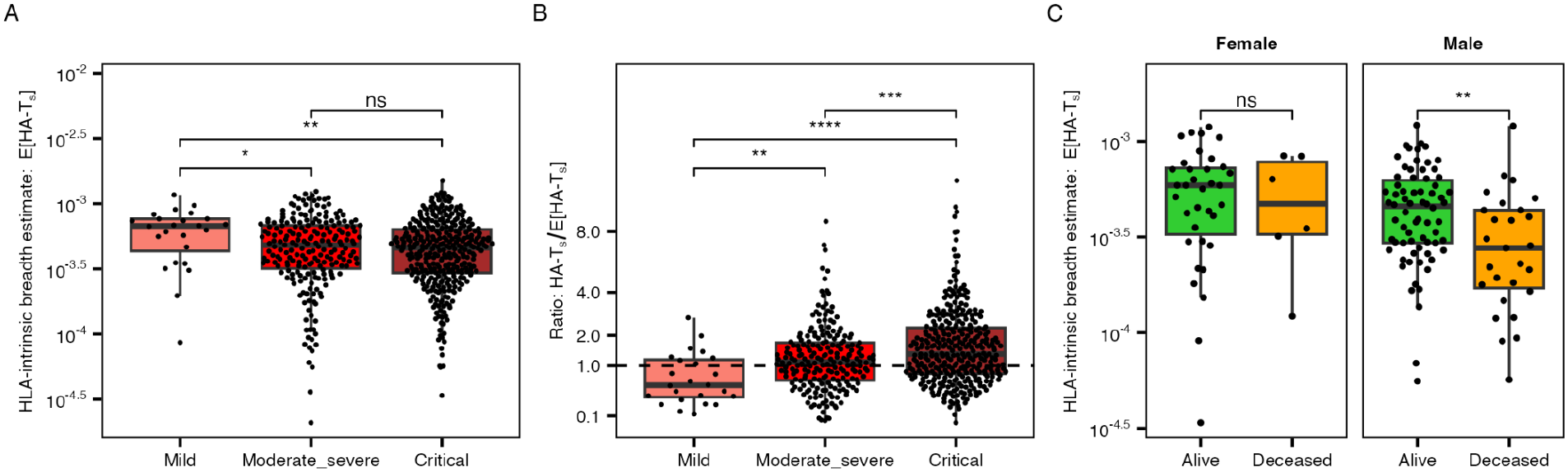
HLA-intrinsic breadth may be inversely associated with severe disease risk. **(A)** Distribution of HLA-intrinsic breadth estimates. Estimated HLA-intrinsic breadth (E[HA-T_S_]) across severity groups. Data shown for repertoire collected 7-45 days from symptom onset. **(B)** Ratio of observed Day 14 breadth to the estimated breadth (HA-T_S_ / E[HA-T_S_]) by severity group. (**C**) Expected HLA-intrinsic breadth in ICU survivors versus non-survivors (Delmonte et al. 2024 study). ****, p < 0.0001; ***, p <0.001; **, p <0.01; *, p < 0.05; ns, not significant.

Despite this clear evidence for antigen exposure to drive observable T_S_ after infection, we reasoned that a low E[HA-T_S,HLA_] might predispose SARS-CoV-2 naive individuals to a poor initial T cell response at baseline, compromising immunological control. Because acute phase repertoires (direct measures of HA-T_S_ after infection) reflect sample timing and intensity of disease course, they therefore are suboptimal for predicting risk. However, we hypothesized that the expected HLA-intrinsic breadth (E[HA-T_S,HLA_]), which is based on genotype information alone, might reflect early T cell response capacity. Consistent with this, E[HA-T_S,HLA_] among men admitted to the ICU, was markedly lower in fatal compared to non-fatal cases (**Fig. 4**C). This relationship between expected HLA-intrinsic breadth and mortality was not observed in women (**Fig. 4**C), which may have a biological basis given prior observations that poor early T cell responses were differentially linked to severe disease trajectories in male versus female patients ^33^.

## Discussion

In this study, we developed an HLA-aware catalog of spike-specific TCRs to quantify the diversity of individual T cell responses to SARS-CoV-2 following infection and vaccination. The post-vaccination HA-T_S_ breadth scores of individuals in the COV-BOOST study were associated with risk of symptomatic COVID-19, with above-median scores associated with approximately 3-fold lower risk independent of measured T cell function (ELISpot) and neutralizing antibody titer. The catalog curation approach integrated TCRs with (i) experimentally validated specificity, or (ii) strong statistical associations across large cohorts with spike antigen exposure, conditioning to individuals with a predicted restricting HLA allele. The resulting catalog contains thousands of TCRβ sequences recruited by HLA-A, HLA-B, HLA-C, HLA-DR, HLA-DQAB, and HLA-DPAB variants, which can be a resource to future studies of SARS-CoV-2 immunity. Further investigation demonstrated that in addition to estimation of observed T_S_ breadth scores, the catalog enables estimation of an individual’s underlying capacity for spike-reactive T cell breadth based on their HLA genotype alone, generating hypotheses about the role of aggregate HLA variation in T cell mediated immunity.

The identification of reliable T-cell immune correlates of protection remains central to evaluating and optimizing next-generation vaccines and evaluating risk-level among populations with partial-T cell immunodeficiencies. Although neutralizing antibody titers have served as valuable surrogates of protection against SARS-CoV-2 and other pathogens ^34^, they capture only one dimension of adaptive immunity. T cells provide complementary protection against symptomatic and severe disease via cytotoxic elimination of infected cells and by supporting high-quality antibody affinity maturation ^35^. Moreover, population level viral escape from T cell recognition is expected to occur more slowly than escape from neutralizing antibodies ^36,37^. As viral evolution undermines the predictive value of pseudoneutralization assays against prior circulating strains, it is critical to determine whether blood-based T cell signatures can also serve as correlates of protection.

In this study, models incorporating HA-T_S,_ or E[HA-T_S,HLA_] predicted the risk of symptomatic infection after booster vaccination more accurately than Day 28 NT_50_ antibody measurements alone. However, this comparison is inherently constrained by the timing of antibody sampling. While the available Day 28 NT_50_ measurement may represent peak neutralization titers against ancestral strains, it may not fully capture antibody durability or cross-neutralization potency against the circulating Delta and Omicron variants. Thus, the association between HA-T_S_ breadth and lower symptomatic infection risk may reflect a direct, cytotoxic layer of T cell-mediated protection. Alternatively, HA-T_S_ breadth might also serve as a second-order biomarker for the quality and persistence of the humoral response that is not fully reflected in the Day 28 NT_50_.

Recent data highlight that decay rates of anti-S antibody vary between individuals ^38^; for example, Park et al. (2025) found that the risk of breakthrough infection was actually highest among individuals with high initial post-booster anti-S IgG titers but rapid antibody decay kinetics, rendering single-point anti-S IgG measurements insufficient to predict risk ^39^. Interestingly, while anti-S IgG and ELISpot metrics measured after vaccination failed to predict infection outcome in that study, Park and colleagues noted that anti-S IgA levels on Day 28 did correlate with protection. Thus, if post-boost SARS-CoV-2 S reactive T cell diversity reflects a well coordinated immune response that supports antibody class switching, affinity maturation, or sustained antibody production, then the protection we observed after vaccination could – at least in part – be antibody-mediated, yet statistically linked to higher levels of HA-T_S_ cell breadth post-boost. Weighing the roles of these two feasible mechanisms of protection will therefore require longitudinal antibody profiling alongside T cell sequencing to test whether post-boost acquired T_S_ cell breadth is linked with differential antibody quality or decay kinetics.

A functional measure of T cell response (ELISpot), which was correlated with HA-T_S,CD8+_ breadth but not HA-T_S,CD4+_ breadth in this study, also predicted a reduction in symptomatic infection risk after booster vaccination. This aligns with direct protection from enhanced cellular mediated immunity, since persons who can mount robust post-infection effector T cell response tend to have better disease outcomes ^17,33^). Nonetheless, the magnitude of initial Spike-specific T cell IFN-γ response may only partially predict the durability and polyfunctionality of T cell memory. Additional signs of a robust spike-reactive clonal diversity, captured by TCR sequencing, could encompass a wider range of receptor affinities beneficial for T cell survival in germinal centers or sustained tissue-resident immunity, consistent with a prior study showing that SARS-CoV-2 T cell clonal diversity, rather than individual clonotype abundance in blood, better predicted persistence of cellular memory ^40^.

The primary study population in the COV-BOOST trial was drawn from multiple sites in the United Kingdom, enriched for so far well studied HLA alleles (e.g., A*02:01, A*03:01, B*08:01), which increases the chance that the SARS-CoV-2 S reactive catalog developed in this study matched vaccine elicited receptors. Although this cohort was not large enough to definitively assess whether risk of symptomatic breakthrough infection varied due to carriership of single HLA variants, it was notable that fewer symptomatic infections were observed in individuals carrying HLA-A*03:01, the second best represented class I allele in the newly developed HLA SARS-CoV-2 S catalog, with over 400 reference receptors. The robust contribution of A*03:01-linked receptors to E[HA-T_S_] breadth is consistent with the hypothesis that specific HLA alleles that activate diverse vaccine acquired CD8^+^ T cells may enhance effective immunological control ^40–42^. Other recent studies have shown A*03:01 carriership may be associated with improved humoral response to vaccination ^43^, reduced risk of breakthrough infections ^44^ and lower rate of long COVID-19 ^45^. We also observed that fewer symptomatic infections occurred in participants with the HLA-DQA*0102/HLA-DQB*0602 haplotype, which was also well represented in the newly developed SARS-CoV-2 catalog. The HLA-DQB1*06 allele has been previously linked to robust antibody responses against the Spike RBD after vaccination in the COV001 and COV002 trials, and HLA-DQB1*06 carriership was previously associated with lower risk of breakthrough infection after primary vaccination in a UK based study population ^46^.

The methodology employed in this study to generate an HLA-linked SARS-CoV-2 spike catalog integrated data from multiple data sets benefited from unprecedented immunological data collection that occurred during the SAR-CoV-2 pandemic. In order to generate similarly effective reference sets for other respiratory pathogens (e.g., Influenza, RSV, or *Mycobacterium tuberculosis*) it is likely that large scale data collection and sharing are needed to provide the foundation for robust and portable TCRseq-based biomarker development. Recently developed tools for HLA imputation directly from TCR repertoire data ^24,47,48^ and exposure-linked TCR clustering graph algorithms ^49,50^ can facilitate the fusion of HLA-labeled and unlabeled data and can help derive TCR-HLA-immunogen associations needed to denoise partial information contained in single chain TCR sequences ^51^ and uncover blood based signals of cellular mediated immunity that could improve durability and population coverage of future vaccines.

## Materials and Methods

### Cohort and Participant Selection

COV-BOOST was a multicentre, randomised, phase 2 trial evaluating third-dose booster vaccination against COVID-19, with a prespecified subgroup undergoing detailed immunological assessments. We utilized samples collected as part of this trial which was conducted at 18 sites across the UK, encompassing both community and secondary-care settings. The trial was reviewed and approved by the South Central–Berkshire Research Ethics Committee and the Medicines and Healthcare products Regulatory Agency. The trial was registered with EudraCT (2021-002175-19) and the Integrated Research Application System (IRAS 299180; REC reference 21/SC/0171).

Participants had previously received two doses of either AstraZeneca ChAdOx1 nCoV-19 (ChAd) or BioNTech BNT162b2 (BNT) at least 84 days prior to the time of enrolment ^19,20^. In this study participant samples were considered if they received one of the following booster vaccines: AstraZeneca ChAdOx1 nCoV-19 (ChAd), Janssen Ad26.COV2.S (Ad26), mRNA BioNTech BNT162b2 (BNT), mRNA Moderna mRNA-1273 (MOD), or protein subunit Novavax NVX-CoV2373 (NVX). Based on sample availability, we analyzed a set of participants with paired Day 0 and Day 14 PBMC samples for TCR sequencing and functional assays. Cases were determined by a positive SARS-CoV-2 test (e.g.,an antigen detection or nucleic acid amplification test) starting 1 week after vaccination. Non-case controls were participants with no evidence of symptomatic SARS-CoV-2 infection through the end of April 2022. Based on sample availability, controls were selected from eligible non-cases in proportion to the treatment assignment of the cases. From available samples, one male and one female non-case participant of similar age was selected for every case. After sample processing, the final dataset comprised 120 participants (43 cases and 77 controls) with matched Day 0 and Day 14 TCR repertoires. Participant HLA information was not considered in sample selection.

### Construction of an HLA-linked SARS-CoV-2 Spike-reactive TCR Catalog

To construct a large catalog of HLA-linked SARS-CoV-2 Spike-reactive T cell receptors (TCRs), we integrated prior public data. We considered evidence from cohort-wide statistical enrichment testing, longitudinal vaccination studies, and previously published experimental validation datasets. Our objective was to maximize sensitivity for identifying candidate Spike-reactive clonotypes from bulk single-chain TCRβ data while preserving information regarding the strength evidence for TCRβ-antigen-specificity through sequence-level confidence weighting.

### HLA-linked SARS-CoV-2-associated TCRs

To identify novel HLA-associated TCRs induced by SARS-CoV-2 infection, we analyzed 1,355 TCR repertoires previously collected from individuals with acute and convalescent COVID-19 ^23,52^. As HLA genotypes were not available for these repertoires, common HLA alleles were imputed directly from the TCR repertoire features using previously described methods for high-confidence HLA genotype inference with TCR2HLA^24^.

We then evaluated public TCRs within these post-infection repertoires for associations with individual HLA alleles. Each TCRβ receptor was represented by its TRBV-gene family (e.g., V05, V01) and its complete CDR3β amino acid (AA) sequences. For each common imputable allele (e.g. A*01:01) or haplotype (e.g. DQA1*05:01/DQB1*02:01), we tested whether a given TCRβ or its edit-1 AA mutational variant was enriched among carriers relative to non-carriers. This analysis identified numerous HLA-associated TCRs that were distinct from previously identified HLA-linked reference receptors – likely associated with other common viral exposures ^24^ – yielding a catalog of putative SARS-CoV-2-associated clonotypes linked to specific HLA alleles. If a receptor sequence (TRBV-family-CDR3(AA) was strongly linked to multiple alleles or haplotypes, we retained all possible associations for downstream detection.

### Identification of Putative SARS-CoV-2-Spike-reactive Clonotypes

Because the post-infection repertoires contained TCRs targeting multiple SARS-CoV-2 antigens, including likely immunodominant responses against the nucleocapsid and ORF1 proteins, we next sought to identify receptors likely reactive to the S protein found in current vaccines. Candidate HLA-linked TCRs were evaluated using paired pre- and post-vaccination repertoires from three previously published cohorts of mRNA vaccine recipients ^26,41,42^, the COV-BOOST cohort, and a separate COV002 cohort. Post-vaccine samples were only considered if they were collected within 7-45 days after two-dose primary or booster vaccination.

For each candidate TCR, we assessed enrichment among individuals carrying the predicted HLA-allele following vaccination. A sequence was retained as putatively Spike-reactive if it was detected more frequently in HLA-matched post-vs. pre-vaccination repertoires. Because relatively few vaccinated participants possessed each inferred HLA allele, we intentionally applied permissive enrichment criteria to maximize sensitivity and retain potentially informative sequences (odds ratio > 1; Fisher’s exact test p < 0.5).

### Assignment of TCRβ-level Confidence Weights

To estimate the strength of evidence supporting each putative Spike-reactive TCR, we compared the frequency of each candidate clonotype between HLA-matched post-vaccination repertoires and pre-2019 SARS-CoV-2-naïve repertoires.Sequences with odds ratios greater than one were retained. We then transformed odds ratios using Yule’s Q statistic:

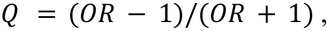

which bounds enrichment estimates between 0 and 1. The resulting Q value was assigned as a sequence-level confidence weight, with larger values, closer to 1, indicating stronger enrichment in post-vaccination repertoires and therefore greater confidence that the detection of that TCRβ clonotype represents S antigen recognition as opposed to sampling noise or an irrelevant TCRα:TCRβ pairing.

### Experimentally Validated Spike-reactive TCRs

To augment the catalog of statistically inferred clonotypes, we incorporated TCR sequences supported by direct experimental evidence of peptide–HLA binding. We curated Spike-reactive clonotypes identified through tetramer or multimer staining assays from previously published studies ^41,53–59^. In addition, we included previously reported HLA-A*03:01-associated Spike-reactive clonotypes identified by activation inducible marker-TCRseq validation through cluster-representative Jurkat peptide stimulation assays ^41^. These experimentally supported sequences were filtered and assigned confidence weights using the same bounded Yule’s Q transformation framework described above.

### Vaccine-responsive HLA-linked Clonotypes

We additionally used longitudinal vaccination studies to identify clonotypes exhibiting evidence of vaccine-induced expansion. Using paired pre- and post-vaccination repertoires from prior studies ^26,41,42^, we quantified clonal expansion of individual TCRs following mRNA vaccination. Clonotypes were retained if they demonstrated both substantial expansion (fold change > 2) and statistical significant change in template counts (FDR-adjusted Fisher’s exact test p < 0.05). To link these expansions to HLA-restricted responses, expanded clonotypes were only retained receptors if they could be assigned to a common HLA allele or haplotype through enrichment testing in the post-infection repertoires (p < 0.01). Confidence weights for kinetically identified clonotypes were assigned using the same Yule’s Q-based framework described above.

### Construction of the Final HLA-linked Spike-reactive TCR Catalog

Thus, the final catalog was assembled by combining evidence from three complementary sources: (i) statistically inferred HLA-linked Spike-reactive clonotypes, (ii) experimentally validated Spike-reactive TCRs, and (iii) vaccine-responsive clonotypes identified through prior longitudinal repertoire analysis. When a clonotype was supported by multiple methods, redundant clones were collapsed into a single record linked to the corresponding HLA allele or haplotype. The resulting catalog contains 7,163 HLA-linked Spike-reactive TCRs annotated with sequence-level confidence weights, which we used to quantify HLA-aware Spike-specific T cell breadth.

### HLA-aware Spike-specific T Cell Breadth (HA-T_S_ Breadth) metric

Antigen specific breadth is a commonly used metric in TCR sequencing analysis to describe the clonotypic diversity of a T cell recognizing an antigen. It is the proportion of clonotypes matching a reference catalog divided by the total unique receptor sequence in the sample. We developed an HLA-aware measure of T cell breadth based on the catalog of HLA-linked Spike-reactive TCRs described above. Unlike conventional breadth metrics that count all diagnostic SARS-CoV-2-reactive clonotypes equally, this approach restricted breadth estimate to only clonotypes linked to HLA alleles carried by the participant and incorporated sequence-level confidence weights (described above).

For each participant, genotyped HLA alleles were compared against the HLA-linked Spike-reactive TCR catalog. Catalog entries corresponding to at least one of the participant’s HLA alleles were considered eligible matches. Participant repertoires were then screened for the presence of eligible catalog clonotypes. Each matched clonotype contributed a sequence-level confidence weight, reflecting the strength of statistical or experimental evidence supporting its Spike specificity. HLA-aware Spike-specific T cell breadth for participant was calculated as:

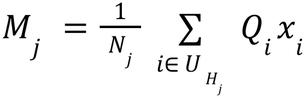

For participant j, **N_j_** is the number of total unique clones in the post-vaccine repertoire sample. **U** is the set of all antigen-associated TCRs in the HLA-CoV2S-TCRβ catalog, and **U_Hj_** denotes the subset associated with the HLA alleles expressed by participant j.

### Inverse Probability Weighted Proportional Hazards Models

Associations between HLA-aware Spike-specific T cell breadth and subsequent risk of symptomatic SARS-CoV-2 infection were evaluated using a calendar-stratified Cox proportional hazards model to account June and November vaccination start months. Inverse probability weights (IPW) were applied based on case status and vaccine platform to account for the two phase case-control sampling design and recover population-level hazard estimates. For IPW weighting, participants were grouped by general prime–boost vaccine modalities: (i) Adenovirus / Adenovirus, (ii). mRNA / mRNA, (ii) Adenovirus / mRNA, (iv) mRNA / Adenovirus, (v). Adenovirus / Protein Subunit, (vi) mRNA / Protein Subunit.

The primary endpoint was time to symptomatic SARS-CoV-2 infection, defined according to the COV-BOOST trial protocol as home antigen test- or PCR-confirmed SARS-CoV-2 infection accompanied by compatible symptoms. Follow-up time was calculated from the date of immunologic sampling until the first documented symptomatic infection or censoring.

The primary immunologic variable was HLA-aware Spike-specific T cell breadth (HA-T_S_). Hazard ratios (HRs) and corresponding 95% confidence intervals were estimated using IPW stratified Cox proportional hazards regression. To evaluate the relative and independent contributions of cellular and humoral immunity, we compared models incorporating HA-T_S_ Breadth, pseudovirus neutralization assay (PNA) neutralization titer 50 (NT50), and IFN-γ ELISpot responses measured at Day 14. NT50 represents potential antibody-mediated protection, whereas HA-T_S_ Breadth and ELISpot capture complementary aspects of cellular immunity, By comparing nested models containing individual and combined immune correlates, we evaluated whether associations between HLA-aware T cell breadth and infection risk persisted after accounting for humoral immunity and conventional functional T cell response magnitude. Analyses were conducted in R version 4.4.3 using the survival package (version 3.8).

We evaluated the following models:

**Model 1 (HA-T_S_ Breadth Model Categorical)**

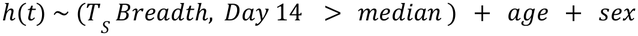

**Model 2 (HA-T_S_ Breadth Model Continuous)**

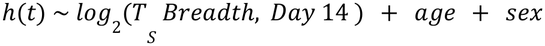

**Model 3 (ELISpot Only Model)**

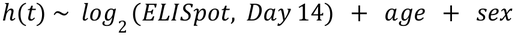

**Model 4 (Antibody Only Model)**

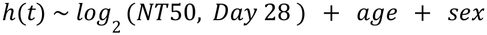

**Model 5 (Combined Correlates Model with Post-Boost Antibody))**

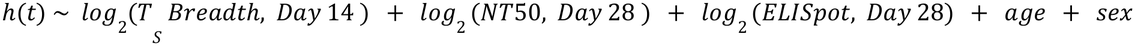

**Model 6 (Combined Immune Correlates Model with Pre-Boost Antibody)**

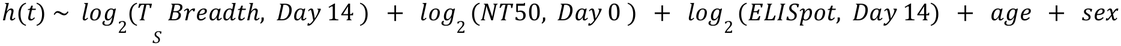

### Sensitivity analyses addressing sequencing-recovery imbalance

We considered whether non-random processing of case and control samples across DNA extraction batches could have biased the results. Because average DNA yield was higher in earlier batches, which disproportionately contained samples from COVID-19 cases (Extended Data **Fig. 5A**), differences in recoverable TCR diversity could potentially confound comparisons of HA-T_S_ breadth between cases and matched controls. To reduce this potential bias, each sample’s repertoire was resampled with replacement to a uniform depth of 100,000 productive templates before computing HA-T[S] breadth (Extended Data **Fig. 5**B). Twenty-two samples (18%) yielded fewer than 100,000 productive templates and were designated low-recovery. Three sensitivity analyses were performed to assess the influence of these samples on the estimated association between HA-TS breadth and risk of symptomatic breakthrough infection:

**Sensitivity Analysis 1:** Low-recovery samples were excluded and the primary Cox model refit in the remaining 98 participants (Extended Data **Fig. 5**C,D model 2),

**Sensitivity Analysis 2:** Imputation of low-recovery values. All participants were retained and Day 14 HA-T_S_ breadth was imputed for low-recovery samples. A linear regression of Day 14 on Day 0 log2(HA-T_S_) breadth was fit in the 98 samples with at least 100,000 productive templates, excluding two near-zero Day 0 breadth values, and used to predict Day 14 values for the low-recovery samples (Extended Data **Fig. 5**F, G). Observed values were retained for all other participants. Imputation was supported by a positive correlation between timepoints in the retained samples (Pearson r = 0.58, p < 0.0001). Each of the three model sets was refit with an additional covariate for inverse repertoire richness, log2(1/R), where R is the number of unique TCRs recovered after downsampling (Extended Data **Fig. 5** models 1A, 2A, 3A), and separately with log2(NT50) (Extended Data **Fig. 5** models 1B, 2B, 3B). All models were stratified, inverse-probability-weighted Cox proportional hazards models adjusted for age and sex, matching the specification of the primary analysis.

### Rank-based Comparisons of Noncases and Cases

To test whether the distributions of biomarkers differed between cases and non-cases, we performed Wilcoxon rank-sum tests (R version 4.4.3)

### Estimation of Expected HLA-intrinsic Breadth

To separate the contribution of HLA genotype from realized T cell responses, we estimated an expected HLA-intrinsic breadth (E[T_S, HLA_]), using only each participant’s HLA genotype. Using an independent cohort of mRNA-vaccinated individuals ^26^, we constructed allele-specific breadth distributions and performed Monte Carlo sampling across the class I and class II alleles carried by each participant to generate a genotype-specific distribution of expected breadth values (Extended Data **Fig. 8**). The median of each participant’s sampled distribution was used as the participant’s expected HLA-intrinsic breadth estimates (E[HA-T_S_]).

We additionally calculated two normalized measures of repertoire performance relative to this genetic baseline. The residual breadth ratio,

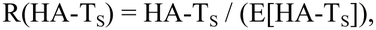

which quantifies observed breadth relative to genotype-derived expectation. We also calculated a breadth quantile score, Q(HA-T_S_) defined as the percentile rank of the observed breadth within the participant’s simulated genotype-specific breadth distribution. These metrics were subsequently evaluated as predictors of infection risk. We compared levels of E[HA-T_S, HLA_], R(HA-T_S_Q), and Q(HA-T_S_) between cases and controls to assess whether infection risk was associated with HLA-intrinsic breadth potential, repertoire performance relative to genetic expectation, or both.

### HLA-Genotyping

Genomic DNA was extracted from peripheral blood mononuclear cells (PBMCs) samples collected at Day 0, Day 14, or Day 28 post vaccination from each unique study participant. DNA extractions were carried out using the QIAamp DNA Blood Mini Kit (Cat. No. 51106; QIAGEN, Venlo, The Netherlands), according to the manufacturer’s instructions. Samples yielding >3.5 µg of total DNA at a concentration of >100 ng/µL were submitted to Histogenetics (Ossining, NY, USA) for HLA genotyping. Prior to sequencing, DNA samples underwent quality control assessment. HLA genotyping was performed using Illumina MiSeq/NovaSeq sequencing to characterize the HLA-A, HLA-B, HLA-C, HLA-DRB1, HLA-DRB3/DRB4/DRB5 (DRB345), HLA-DQB1, HLA-DQA1, HLA-DPB1, and HLA-DPA1 loci at 1X high-resolution level.

### TCR Sequencing of COV-BOOST Samples

For the COV-BOOST study, genomic DNA was isolated (as described for HLA-genotyping) from four million, T-cell Xtend (Catalog number: TTK610, Oxford Immunotec, Abingdon, UK) isolated PBMCs, sourced from whole blood samples (Southampton, UK). Multiplex PCR amplification and high-throughput sequencing of TRBV gene segments and CDR3β regions of the human T-cell receptor β (TCRβ) locus were performed using the standard ImmunoSEQ protocol (Adaptive Biotechnologies, Seattle, WA). Six participants were excluded from downstream analysis because their sample sets possessed insufficient starting DNA material (flagged by Adaptive Biotechnologies’ laboratory) or lacked minimal clonal overlap between timepoints, suggesting a sample labeling error. To reduce variability due to differences in DNA extraction yield and sequencing depth, TCR repertoires were resampled with replacement to a uniform depth of 100,000 templates per sample prior to downstream analyses.

### Previously Developed TCR-based SARS-CoV-2 S Metrics

To compare among HLA-aware Spike-specific breadth and previously established HLA-agnostics approaches, we calculated several published SARS-CoV-2-specific TCR sequencing metrics from each repertoire. These included Adaptive Biotechnologies’ proprietary Spike breadth metric, MIRA-derived SARS-CoV-2 breadth, and the Spike V3 classifier score ^21,22^.

The Spike breadth metric was defined as the proportion of unique TCR clonotypes matching previously identified Spike-associated public TCR sequences.The Spike V3 score represents a logistic regression based classifier-derived measure based on Spike TCRs predictive of SARS-COV-2 exposure. MIRA breadth was calculated using experimentally mapped SARS-CoV-2-reactive TCRs identified through Multiplex Identification of T-cell Receptor Antigen Specificity (MIRA) assays ^23^ but not necessarily part of the diagnostic set of enhanced sequences ^21^. All metrics were evaluated alongside HLA-aware Spike-specific breadth in case vs non-case comparisons. Previously developed adaptive SARS-CoV-2 S TCR metrics were computed on the exact same resampled repertoires as were used in the primary analysis.

### Interferon-γ ELISpot

For cellular immunology readouts, PBMC samples were collected from nine sites based on logistical reasons (i.e. proximity to the external laboratory). Interferon-γ (IFN-γ)-secreting T cells were quantified by stimulating PBMCs from study participants with 15-mer peptides spanning the Wuhan-Hu-1 protein sequence (YP_009724390.1), with an 11-amino-acid overlap between consecutive peptides. IFN-γ-secreting T cells were detected using a modified T-SPOT Discovery assay (Oxford Immunotec, Abingdon, UK). Samples were processed within 32 hours of venepuncture, with the T-Cell Xtend reagent added to remove red blood cells, and granulocytes and extend PBMC viability during sample processing. T-cell responses were reported as spot-forming cells (SFCs) per 250,000 PBMCs. The lower limit of detection was 1 SFC per 250,000 PBMCs. Results were multiplied by four to express T-cell frequencies as SFCs per 1 × 10^6^ PBMCs.

### Serological assays

Serum samples collected at Day 0 and Day 28 post-vaccination were analysed at Nexelis (Laval, QC, Canada) to determine concentrations of anti-spike IgG antibodies against the Wuhan-Hu-1 strain of SARS-CoV-2 using an enzyme-linked immunosorbent assay (ELISA). Results were reported as ELISA laboratory units per millilitre (ELU/mL). Serum samples were analysed at the UK Health Security Agency (UKHSA), Porton Down, UK, to determine the 50% virus-neutralising antibody titre (NT_50_) against live Wuhan-strain SARS-CoV-2 using a microneutralisation assay. All assays were performed in at least duplicate. For the participants with complete pre- and post-booster TCRβ data (n=120), we imputed a small amount of missing functional assay data to preserve the dataset for multivariate models: Day 28 ELIspot (n = 1), Day 28 antibody NT50 (n = 3), and Day 0 NT50 (n = 4). Imputation was performed using a k-nearest neighbors (kNN) algorithm via the VIM package in R. Missing values were estimated based on the 9 nearest neighbors (k=9) using age and vaccine platform to identify most similar profiles.

## Manuscript information

### Author contributions

Conceptualization: KMB, KH, TL, MV, AFG

Methodology: KMB, KH, APSM, AFG

Software: KMB

Validation: KMB, KH

Formal Analysis: KMB, KH, AFG

Investigation: KMB, KH, NE, APD, JA, APSM, SNF

Resources: KH, NE, ESF

Data Curation: KH, NE, ESF, XL

Writing – Original

Draft Preparation: KMB, KH

Writing – Review and Editing: KMB, KH, APD, APSM, SNF, TL, MV, AFG

Visualization: KMB

Supervision: TL, MV, AFG

Funding Acquisition: TL, MV, AFG

### Data and Code Availability

The TCR repertoire data generated in this study have been deposited in Zenodo (https://doi.org/10.5281/zenodo.23965835) and will be made publicly available upon publication in a peer-reviewed journal. The analysis code is available at <u>github.com/kmayerb/spike-tcr-atlas</u>. Participant ages in the public dataset are rounded to the nearest decade, so analyses from this data may differ slightly from the reported results.

### AI disclosure

Authors used an LLM (Microsoft Copilot 365) for proofreading portions of the manuscript text. Claude Desktop (Opus 4.8, 2026) was used to review and finalize code for data visualizations during manuscript preparation. All scientific content, citations, and interpretations are the authors’, and the authors assume full responsibility for the accuracy of the analysis and text.

### Competing interests

The University of Oxford entered into a partnership with AstraZeneca to develop the ChAdOx1 nCoV-19 (AZD1222) COVID-19 vaccine. T.L. is named as an inventor on a patent application relating to this SARS-CoV-2 vaccine (PCT/GB2012/000467). S.B. contributed to several studies involving the clinical evaluation of AZD1222. AstraZeneca facilitated and funded the manufacture of the AZD1222 vaccine candidate and provided financial support for the expansion of Oxford-sponsored clinical trials in the UK and Brazil. S.N.F. has received grants from Sanofi, Pfizer, GSK J&J, AstraZeneca, Moderna, BioNTech, and Cidara, and honoraria were paid to his institution (no personal payments) for participation in vaccine advisory boards or symposia from Moderna, Novavax, Pfizer, Sanofi, Astrazeneca, Seqirus and MSD. Other authors have no conflicts to report.

### Funding support

This work was supported by a Gates Foundation grant to Oxford University (INV-042533). KMB, AFG were supported by a grant from the Gates Foundation (INV-027499) and by NIH grant number R01 AI136514. The conclusions and opinions expressed in this work are those of the authors alone and shall not be attributed to the Gates Foundation. Scientific Computing Infrastructure at Fred Hutchinson Cancer Research Center was funded by an ORIP grant S10OD028685.

### NIH public access statement

This work is the result of NIH funding, in whole or in part, and is subject to the NIH Public Access Policy. Through acceptance of this federal funding, the NIH has been given a right to make the work publicly available in PubMed Central.

## Data Availability

The TCR repertoire data generated in this study have been deposited in Zenodo (https://doi.org/10.5281/zenodo.23965835) and will be made publicly available upon publication in a peer-reviewed journal. The analysis code is available at github.com/kmayerb/spike-tcr-atlas. Participant ages in the public dataset are rounded to the nearest decade, so analyses from this data may differ slightly from the reported results.

https://github.com/kmayerb/spike-tcr-atlas

## Acknowledgments

We thank Dr. Thomas Bello for technical assistance computing the Adaptive TCRseq metrics for COV-Boost TCR repertoires. We thank Drs. Anastasia Minervina, Hiroyasu Aoki, Mikhail Pogorelyy, Stefan Schattgen, Phil Bradley, and Paul Thomas for their helpful comments on this work.

---

**Extended Data Fig. 1:**
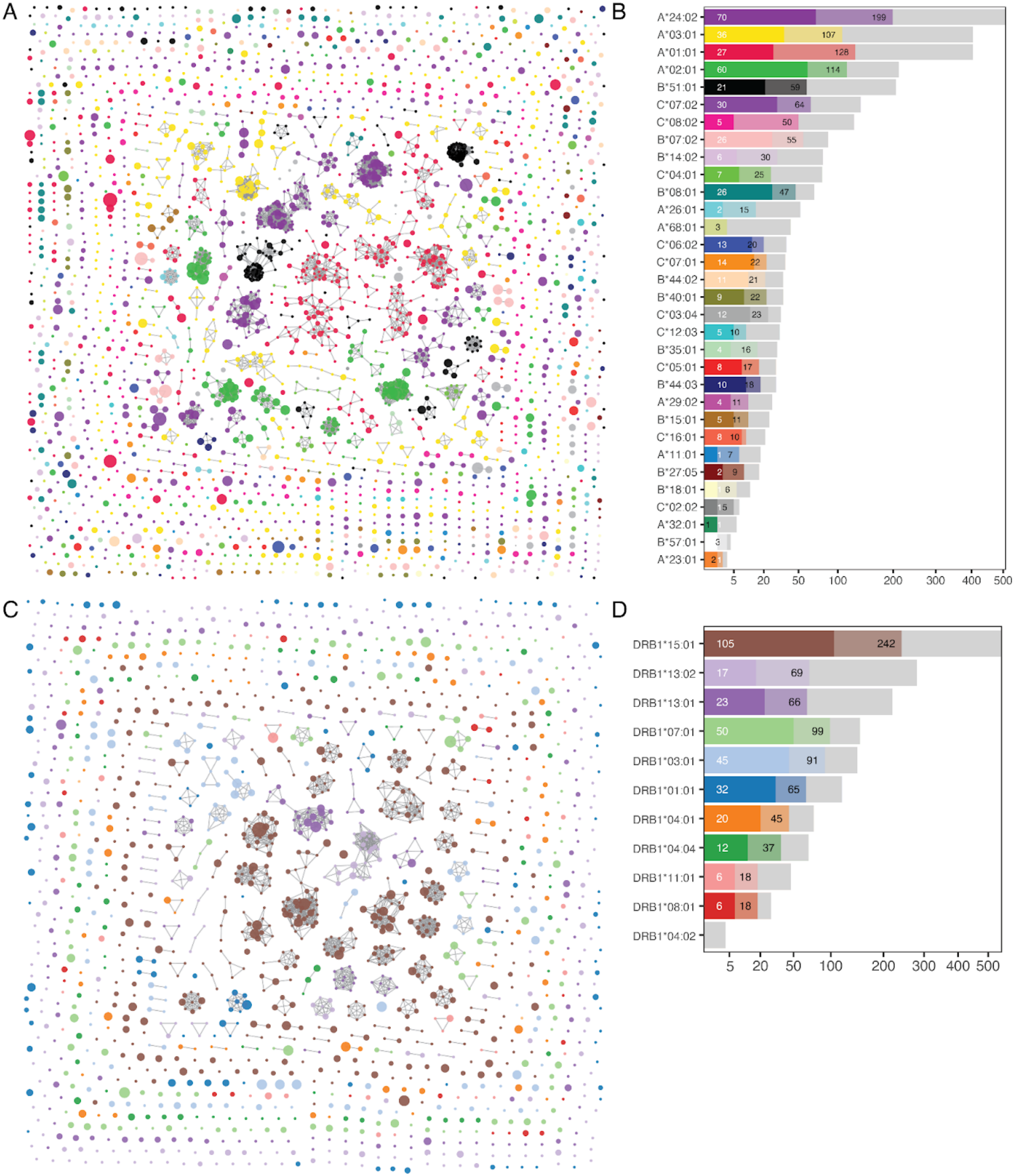
Diverse HLA-associated TCR features are represented in the SARS-CoV-2 catalog. (**A,C**) Sequence similarity network of HLA-associated TCR features from the SARS-CoV-2 spike protein catalog, clustered at a TCRβ distance threshold of < 25 TCRdistance units, retaining edges between nodes with the same predicted HLA association. Node size is scaled to the number of individuals with the predicted HLA genotype in the COV-Boost cohort (n=120) in whom the TCR is detected in post-vaccination (Day 14) TCRβ repertoires (after normalization to 100,000 unique reads per sample). (**B, D**) Number of unique TCRβ detected by specific HLA-A/B/C (B) or *HLA-DRB1 (**D**)* allele association. The horizontal gray bars indicate the total number of HLA-associated features defined in the SARS-CoV-2 catalog. Colored bars represent the recovery of these features within the COV-Boost cohort: light-shaded bars denote features identified in one individual carrying the specified HLA allele, while dark-shaded bars represent “public” features detected in two or more HLA-matched individuals.

**Extended Data Fig. 2:**
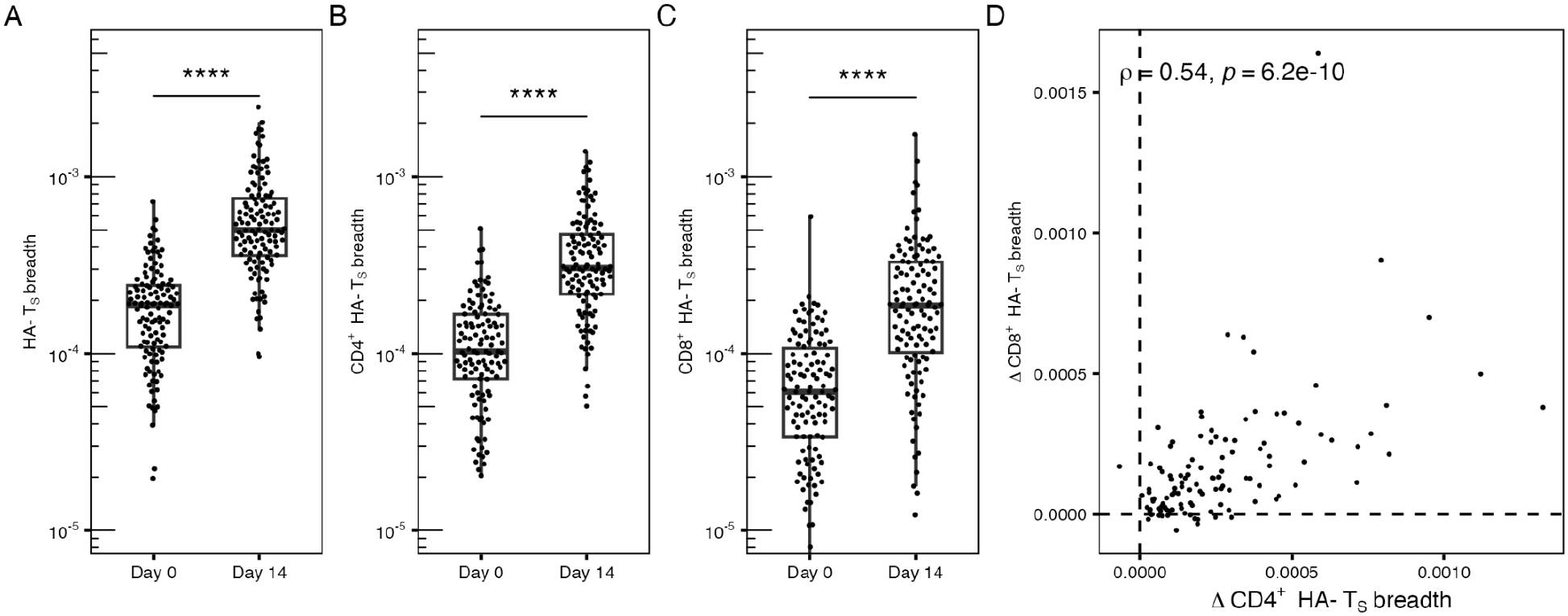
Vaccine-induced expansion of HA-T_S_ breadth across T cell subsets. (**A–C**) Paired analysis of HA-T_S_ breadth at Day 0 (pre-boost) and Day 14 (post-boost) across 120 participants. Plots display the total T_S_ breadth (**A**), CD4^+^ HA-T_S_ breadth (**B**), and CD8^+^ HA-T_S_ breadth (**C**). Statistical significance between timepoints was evaluated using a, Wilcoxon paired signed-rank test (**** indicates p < 0.0001). (**D**) Correlation between the vaccine-induced change (Δ, calculated as Day 14 minus Day 0) in CD4^+^ HA-T_S_ breadth and CD8^+^ HA-T_S_ breadth. ρ indicates the Spearman rank correlation. ****, p < 0.0001; ***, p <0.001; **, p <0.01; *, p < 0.05; ns, not significant.

**Extended Data Fig. 3:**
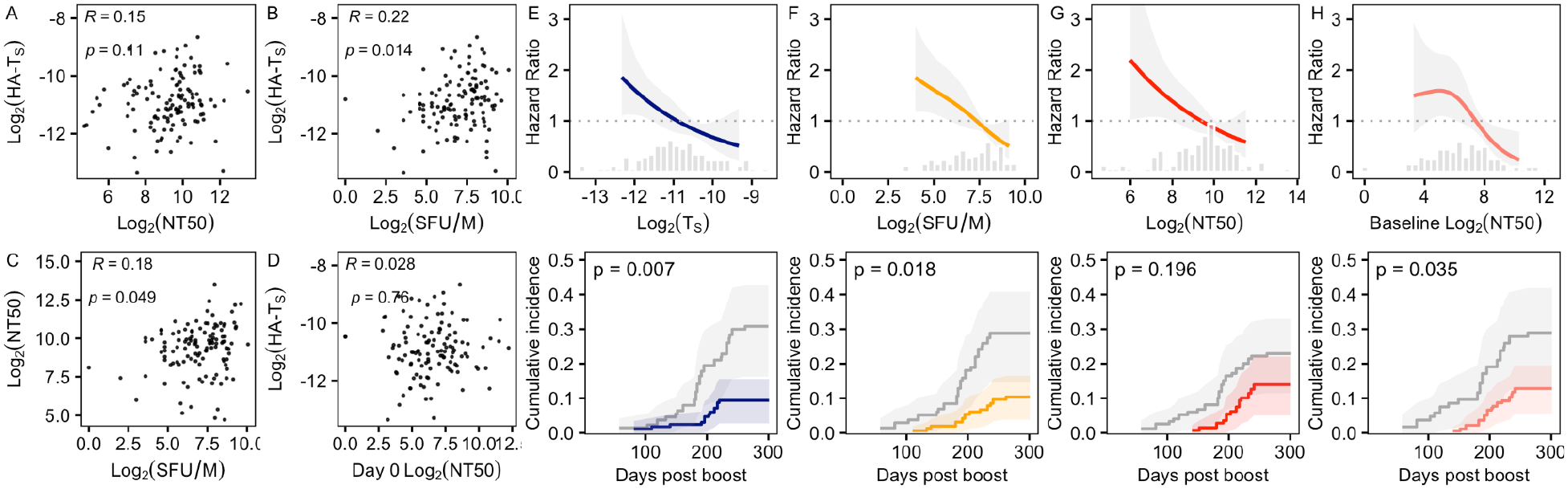
Association of HA-T_S_ with symptomatic SARS-CoV-2 infection and other immune measures. (**A–D)** Correlation analysis of Day 14 HA-T_S_ breadth with alternative immune markers, including ELISpot responses (SFU/M) and Day 28 neutralizing antibody titers (NT_50_). (**E-H**) Hazard ratio plots (top row) showing the estimated relationship between immune metrics (Day 14 HA-T_S_, Day 14 ELISpot, Day 28 NT_50_ and Day 0 NT_50_) and risk of symptomatic SARS-CoV-2 infection after booster vaccination. Models were fit using restricted cubic splines in Cox proportional hazards regression, adjusted for age and sex. Corresponding Kaplan-Meier cumulative incidence curves (bottom row) shown for June vaccination group (n = 90) are stratified by immune marker split at its post-boost, study-wide median value.

**Extended Data Fig. 4:**
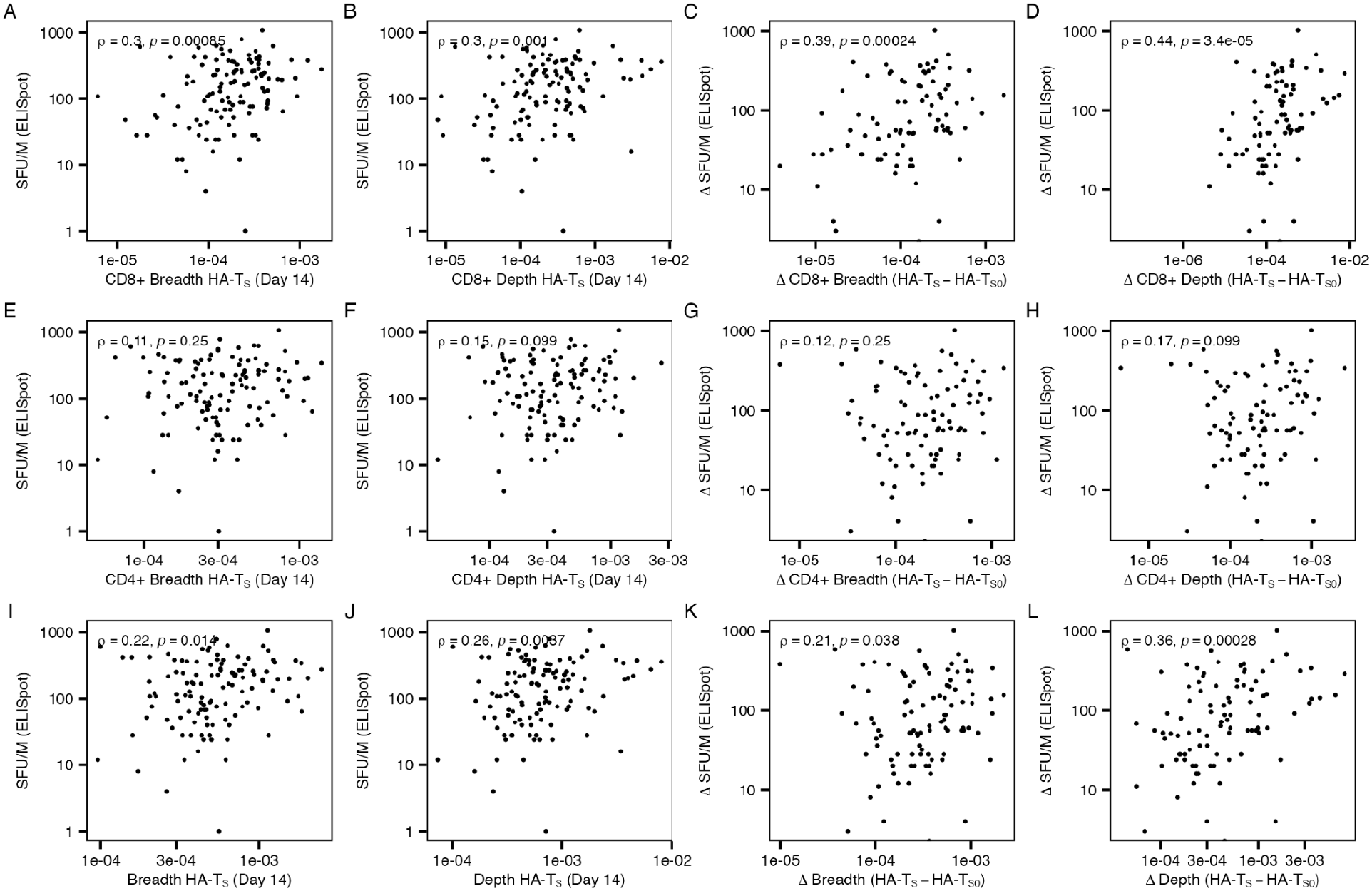
Correlation between ELISpot and HA-T_S_ breadth and depth measures. Each panel shows the per-participant association between an HLA-aware TCRseq-derived measure (x-axis) and functional T cell response magnitude by ELISpot (SFU/M, y-axis) or change in ELISpot between Day 0 and Day 14 in the COV-Boost cohort (n = 120). Columns present, from left to right: Day 14 breadth, Day 14 depth, change in breadth (HA-T_S Day14_ − HA-T_S Day 0_), and change in depth (HA-T_S Day14_ − HA-T_S Day 0_). **(A-D)** Rank correlation between overall ELISpot response versus CD8^+^ HA-Tₛ breadth and depth at Day 14, correlation between change in ELISpot and change in CD8^+^ HA-Tₛ metrics from baseline.**(E-H).** Rank correlation between overall ELISpot response versus CD4^+^ HA-Tₛ breadth and depth at Day 14, correlation between change in ELISpot and change in CD4^+^ HA-Tₛ metrics from baseline. **(I-L)** Total (CD4+ and CD8+ combined) signal. Overall ELISpot response versus overall HA-Tₛ breadth and depth at Day 14 and their change from baseline.

**Extended Data Fig. 5:**
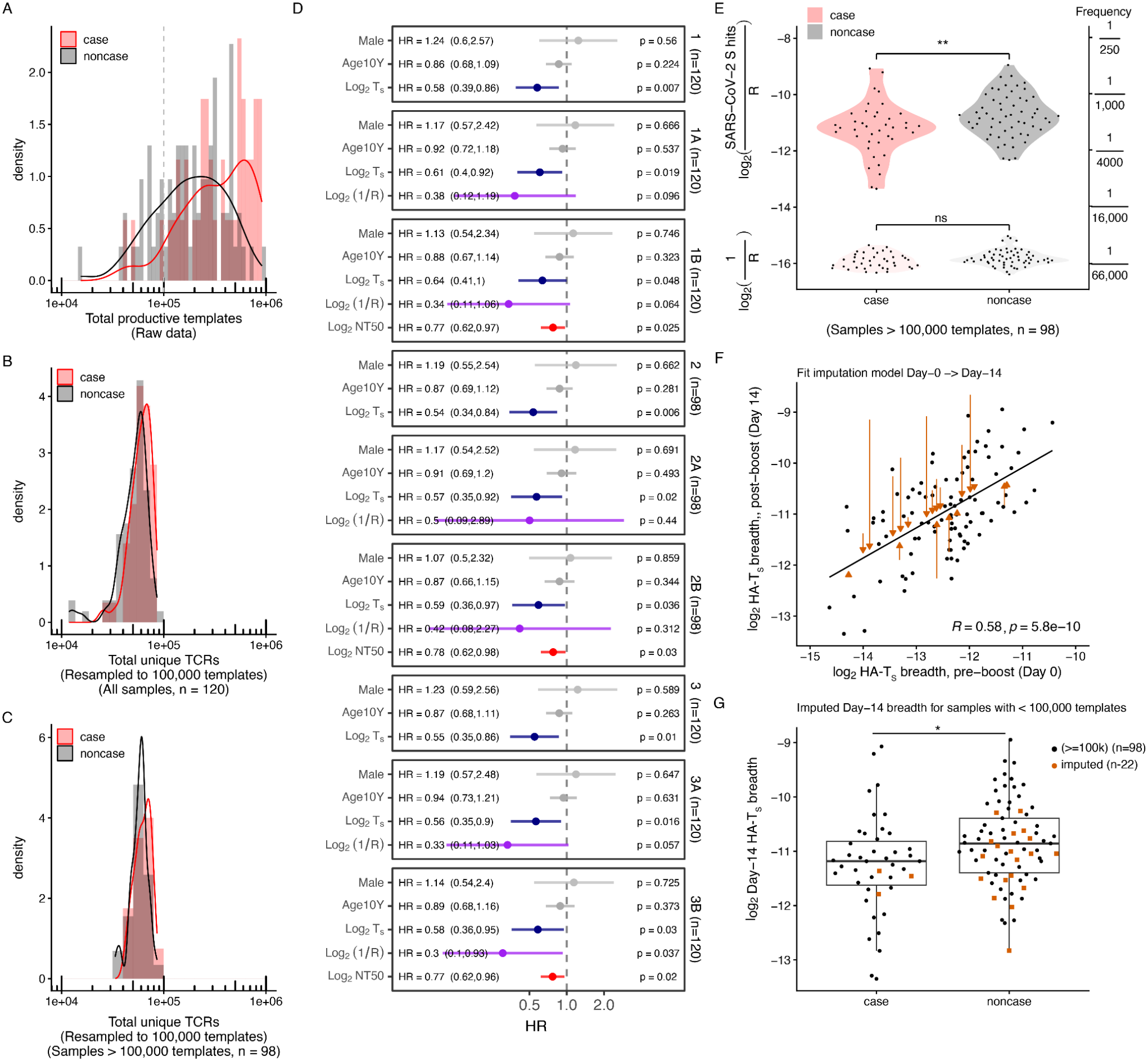
Sensitivity analyses confirm primary results. (**A**) Distribution of total productive templates per sample by breakthrough status. The dashed line marks the 100,000-template threshold used to define low recovery. (**B**) Total unique TCRs after resampling with replacement to 100,000 templates, all samples (n = 120). (**C**) Total unique TCRs after restricted to the 98 samples with ≥ 100,000 productive templates. (**D**) Hazard ratios (95% CI) from stratified, IPW Cox hazard models of symptomatic breakthrough infection. Models 1-1B use all samples with observed Day 14 breadth; models 2–2B exclude low-recovery samples; models 3–3B impute Day 14 breadth for low-recovery samples from a Day 0 regression. The model A variants add inverse richness, log2(1/R), where R is the number of unique TCRs after downsampling; the model B variants add Day 28 log2(NT50) a measure of antibody neutralization. All models adjust for age (per 10 years) and sex and are stratified by vaccination start month. (**E**) Day 14 HA-T[S] breadth (upper) and inverse richness (lower) by breakthrough status among deeply-sequenced samples (n = 98); brackets show two-sided Wilcoxon rank-sum tests (** p < 0.01; ns, not significant). The right axis estimates the inverse-richness scale as unique TCRs per template. (**F**) Day 0 versus Day 14 log2 HA-T[S] breadth in deeply-sequenced samples, with the fitted imputation model (line) and Pearson correlation. Orange arrows run from each low-recovery sample’s observed value to its imputed value. (**G**) Day 14 breadth by breakthrough status after imputation of low recovery samples, (n = 120); two-sided Wilcoxon rank-sum test.

**Extended Data Fig. 6:**
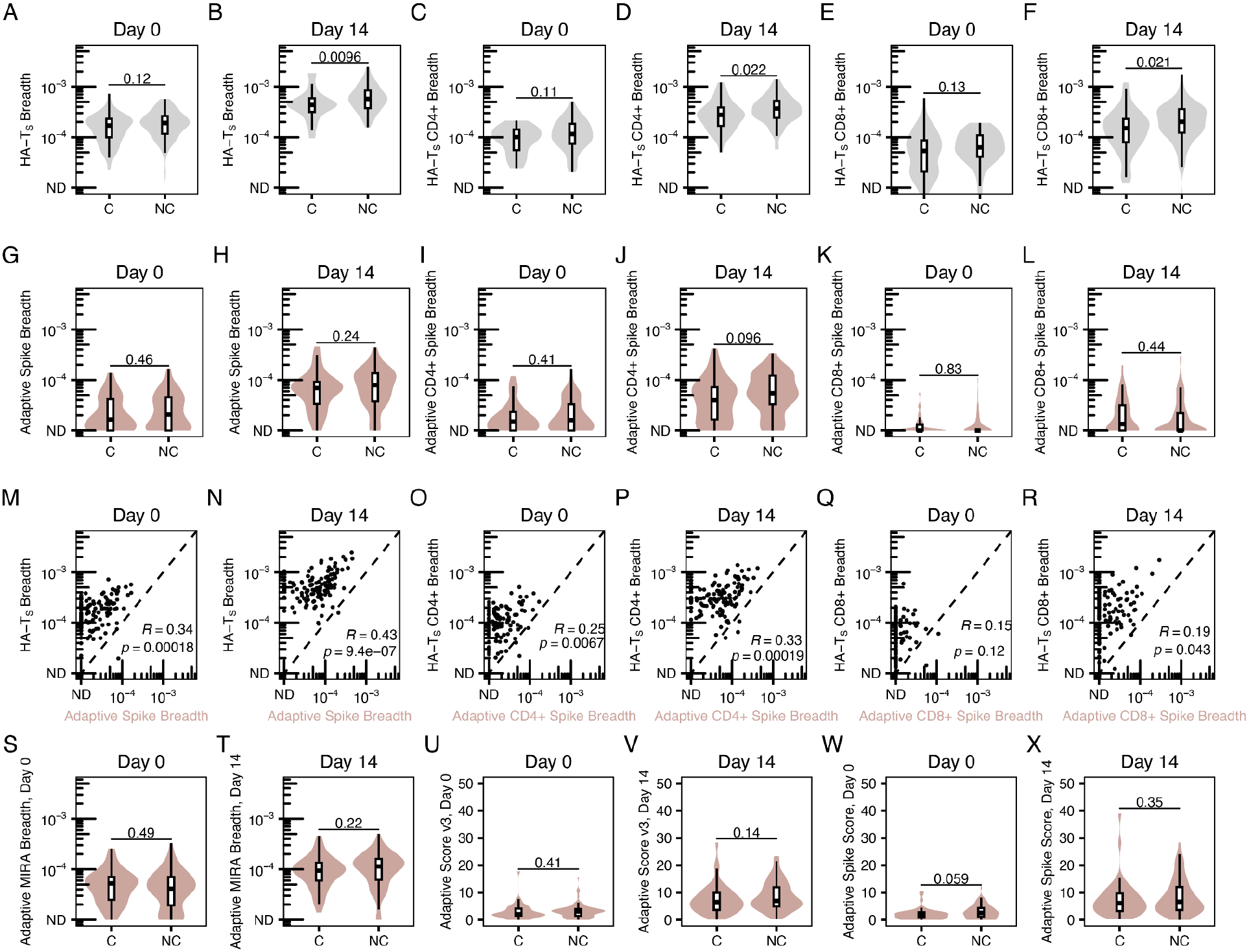
HLA-aware T cell breadth distinguishes breakthrough cases from non-cases more than HLA-agnostic TCRseq-based metrics. (A–F) Primary HLA-aware HA-Tₛ breadth (overall, CD4+, CD8+) in symptomatic breakthrough cases (C) versus non-cases (NC) at Day 0 and Day 14. Violins show the full distribution; boxplots show median and IQR. (G–L) Analogous previously published adaptive (TCRseq) spike metric breadth measures (overall, CD4+, CD8+) at Day 0 and Day 14. (M–R) Per-participant concordance between Adaptive breadth (x-axis) and primary HA-Tₛ breadth (y-axis) for overall, CD4+, and CD8+ compartments at each timepoint; dashed line denotes y = x, and R/p are Spearman correlation. Cases are shown a black filled points. (S–X) Additional Adaptive metrics: MIRA breadth, Score v3, and Spike Score at Day 0 and Day 14. All axes are log₁₀ except the Adaptive scores (U, V, W, X), which are linear; non-detects are plotted at the ND floor. Group comparisons are Wilcoxon rank-sum tests. Adaptive refers to methods previously developed by Adaptive Biotechnologies.

**Extended Data Fig. 7:**
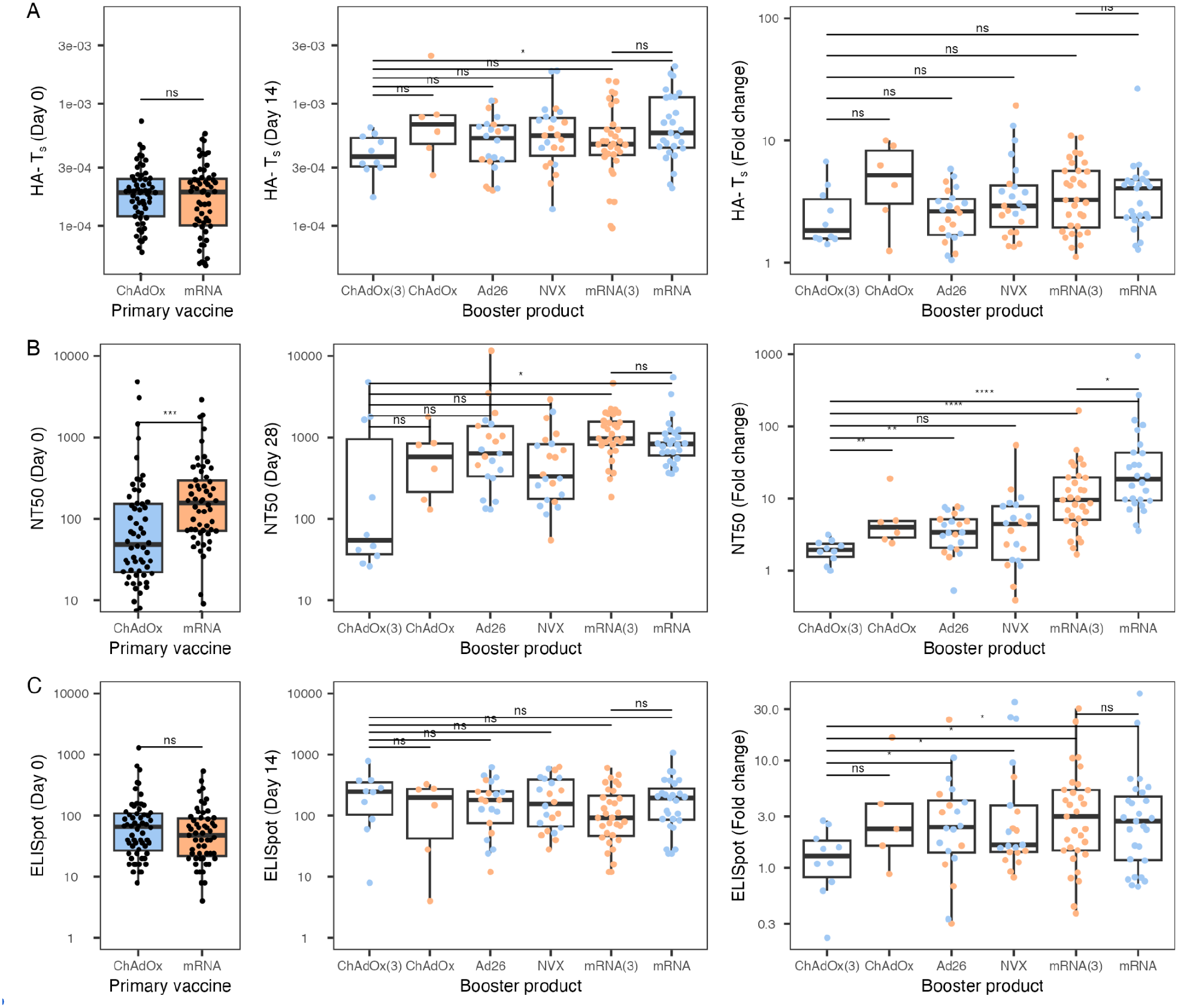
Difference in Antibody and T Cell immune response across prime and booster vaccine modalities. (**A**) HLA-aware T cell clonotypic breadth (HA-T_S_), (**B**) Antibody neutralization (NT50), and (**C**) Spike-specific T cell responses (ELISpot SFU/M) before and after booster vaccination. Baseline (Day 0) levels grouped by primary vaccine category (ChAdOx vs. mRNA). Day 14 responses by booster vaccine type (ChAdOx(3), ChAdOx, Ad26, NVX, mRNA. Fold change from Day 0 to Day 14 by vaccine booster modality. Boxplots show median and IQR with individual participants overlaid; pairwise p-values are Wilcoxon rank-sum. ChAdOx(3) indicates the group receiving homologous ChAd vectored vaccine after primary ChAdOx series. mRNA (3) indicates persons only receiving mRNA vaccines. Colors of the points indicate the primary vaccine type.

**Extended Data Fig. 8:**
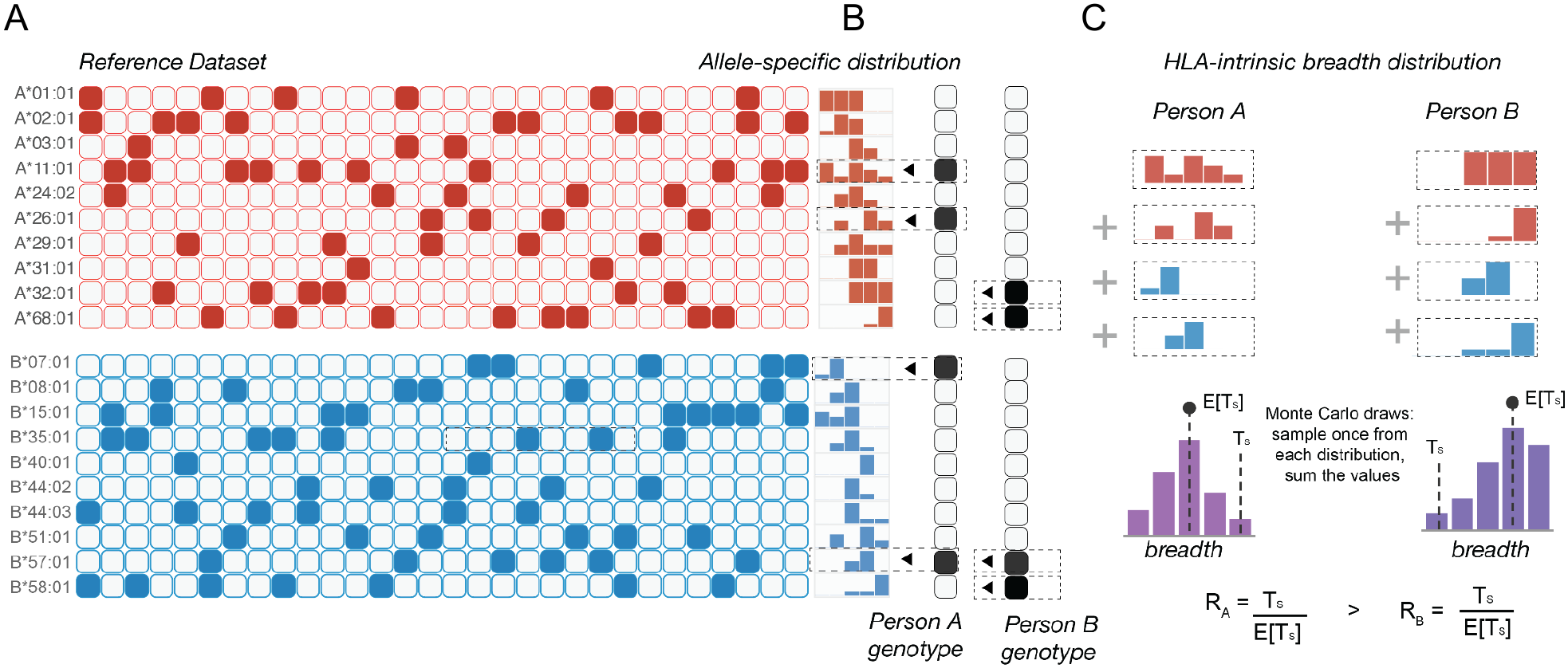
Procedure for estimating expected HLA-intrinsic breadth. Expected HLA-intrinsic breadth, E[Tₛ,HLA], is derived for each participant from their HLA genotype alone. (**A**) In an independent reference cohort of mRNA-vaccinated individuals, allele-specific breadth distributions are constructed for each HLA allele (only HLA-A and HLA-B shown for simplicity). Each row corresponds to one allele; the small histograms at right illustrate the distribution of observed breadth values across reference individuals from another study carrying that allele. (**B**) For each study participant (e.g., person A, person B), the alleles present in their genotype are used to retrieve the corresponding allele-specific breadth distributions. (**C**) Then, a genotype-specific distribution of expected breadth is generated by Monte Carlo sampling — drawing once from each of the participant’s allele distributions and summing the values across their carried class I and class II alleles. The resulting per-participant distribution (purple) yields the expected HLA-intrinsic breadth, E[Tₛ,HLA] (median, point/dashed line), against which the participant’s observed breadth, HA-Tₛ (dashed line), is compared. The residual breadth ratio (R), HA-Tₛ / E[HA-Tₛ], quantifies observed breadth relative to this genotype-derived expectation. Persons A and B illustrate contrasting cases: person A carries alleles conferring lower expected breadth, whereas person B’s genotype yields a higher expectation.

**Extended Data Fig. 9:**
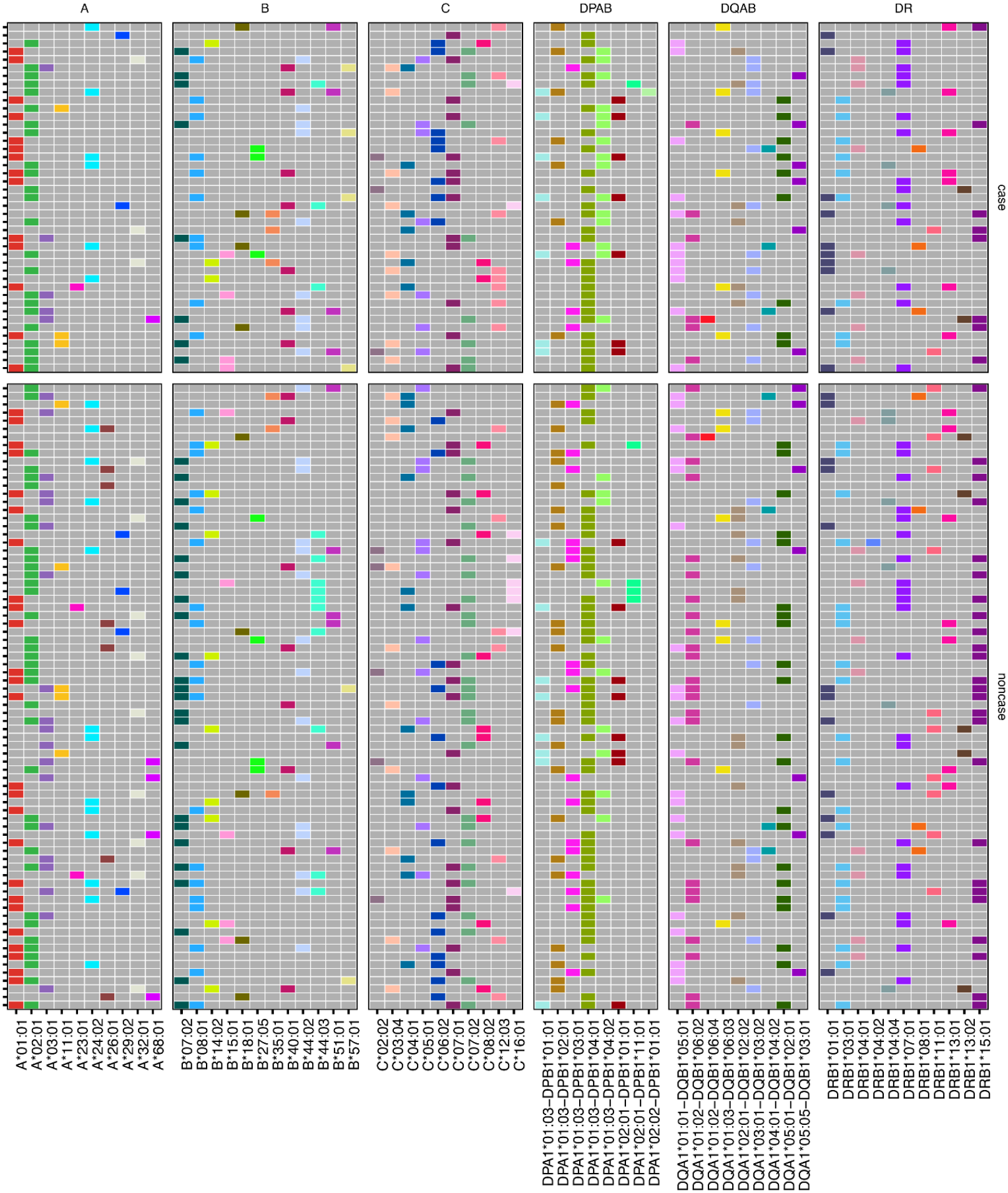
HLA Genotypes in the COV-BOOST cohort by case and noncase status. Participant-level HLA genotypes used in the SARS-CoV-2 Spike-reactive TCR catalog. Rows represent participants; columns show HLA-A, HLA-B, HLA-C, and HLA-DRB1 alleles, as well as HLA-DQ and HLA-DP heterodimers.

## Notes

### Clinical Trial

EudraCT Number: 2021-002175-19

### Author Declarations

The trial was reviewed and approved by the South Central-Berkshire Research Ethics Committee and the Medicines and Healthcare products Regulatory Agency. The trial was registered with EudraCT (2021-002175-19) and the Integrated Research Application System (IRAS 299180; REC reference 21/SC/0171).

